# Host-microbiome determinants of ready-to-use supplemental food efficacy in acute childhood malnutrition

**DOI:** 10.1101/2024.12.12.24318367

**Authors:** Zehra Jamil, Gabriel F. Hanson, Junaid Iqbal, G. Brett Moreau, Najeeha Talat Iqbal, Sheraz Ahmed, Aneeta Hotwani, Furqan Kabir, Fayaz Umrani, Kamran Sadiq, Kumail Ahmed, Indika Mallawaarachchi, Jennie Z. Ma, Fatima Aziz, S Asad Ali, Sean Moore

**Author notes:** These authors have contributed equally to this work (Zehra Jamil, Gabriel F. Hanson).

## Abstract

Ready-to-use supplemental foods (RUSF) are energy-dense meals formulated to prevent and treat moderate and severe childhood acute malnutrition (MAM and SAM) in high-risk settings. Although lifesaving, the degree and durability of weight recovery with RUSF is unpredictable. We examined whether environmental enteric dysfunction (EED) and gut microbiota perturbations are risk factors for RUSF failure in a birth cohort of 416 rural Pakistani children followed for growth, common childhood illnesses, and biomarkers from blood, urine, and stool. Infants who developed wasting (weight-for-length Z score <-2, n=187, 45%) during surveillance received Acha Mum (a chickpea-based RUSF) daily for eight weeks. Machine learning identified seven biomarkers that predicted RUSF response (n=75) vs. non-response (n=112) with 73% accuracy. Remarkably, gut microbiome composition predicted RUSF response with 93% (pre-supplementation) and 98% (post-supplementation) accuracy. Seven outliers whose microbiome falsely predicted positive response experienced extraordinary burdens of inflammation and illness during supplementation. These findings identify gut microbial signatures and biomarkers of gut and systemic inflammation as robust predictors of RUSF response in infants free from intercurrent illness during recovery, setting the stage for predictive models to guide precision use of RUSF and adjunct therapies in undernourished children.

## Introduction

The 2025 WHO Global Nutrition Targets call for dramatic and overdue reductions in the global burden of childhood wasting (acute undernutrition) and stunting (chronic undernutrition) [1]. Persistent health and economic disparities in low- and middle-income countries (LMICs) continue to impede progress toward these goals [2]; hence, the timely application of effective public health interventions as well as an improved understanding of the mechanisms underlying childhood undernutrition in LMICs will be crucial to success [3]. Ready-to-use supplemental foods (RUSFs) have emerged as a key strategy for the prevention and treatment of moderate and severe acute malnutrition (MAM, -2> weight-for-height Z (WLZ)>-3; SAM, WLZ<-3 SD) in community-based settings. Efforts to encourage the local production of RUSF, enhance efficacy and adherence, and reduce gastrointestinal side effects (e.g., vomiting) are on the rise [4]; however, RUSF interventions continue to show inconsistent efficacy and durability for growth recovery [5]. Optimizing RUSF composition, duration, monitoring, and adjunctive therapies to maximize nutritional benefit is currently constrained by a lack of precision tools to identify those children least likely to respond.

Childhood growth faltering in LMICs is closely linked to environmental enteric dysfunction (EED), a subclinical enteropathy characterized by gut and systemic inflammation, intestinal barrier dysfunction, and malabsorption [6]. It is not yet clear to what extent EED impairs the efficacy of RUSF in childhood wasting. Intensive nutritional and WASH (water, sanitation and hygiene) interventions against childhood stunting have so far produced only modest benefits, raising concerns that the derangements in gut structure and function seen in EED undermine these strategies [7]. Recently, causal links between gut microbiota and early childhood undernutrition have been identified, leading to investigations exploring microbiota-directed complementary foods (MDCF) [8, 9]. Although the role of EED in stunting pathways is increasingly appreciated, the degree to which EED impairs the efficacy of RUSF and MDCF is not yet understood.

Building on these observations, we explored whether host-microbiome factors influence RUSF response in acutely undernourished children to address the following questions: 1) To what extent do EED and systemic inflammation undermine the efficacy of RUSF?; 2) Do baseline differences in gut microbiota prior to RUSF administration predict clinical outcomes?; and 3) Do RUSF-driven shifts in gut microbial communities differentiate responders from nonresponders? Here, we report findings from secondary analyses of the Study of Environmental Enteropathy and Malnutrition (SEEM)-Pakistan in which 187 children with recalcitrant wasting received a chickpea-based RUSF. We find that: 1) biomarkers of systemic inflammation, EED, and nutritional status (fecal myeloperoxidase and neopterin; serum prealbumin, glucagon-like peptide-2, and C-reactive protein; and urine claudin-15 and creatinine) collectively predict RUSF response, 2) pre-RUSF microbiome composition predicts RUSF response and 3) RUSF expands the relative abundance of Negativicutes (including *Veillonella* chickpea fermenters) in wasted infants, with responders displaying greater relative increases in Negativicutes and Clostridia, and decreases in Gammaproteobacteria, relative to non-responders.

## Results

### Participant characteristics

We enrolled 416 newborns (3-to-6 months of age) in the rural Matiari district of Sindh, Pakistan. We monitored their weight and length monthly for six months or longer then classified study participants as either well-nourished controls (WLZ ≥0, LAZ ≥-1, n = 51) or undernourished cases (wasted with WLZ ≤-2, n = 365) based on two consecutive months of anthropometry **Fig 1A**). Baseline clinical characteristics for case and control infants for whom complete biomarker and fecal 16S microbiome data were available are summarized in **Table 1**. Maternal and paternal BMI as well as birth anthropometrics were significantly lower in cases vs. controls (P <0.05), differences that became even more pronounced by nine months of age (P <0.05). Breastfeeding patterns were similar between groups over the first six months of life. Cases in whom complete biomarker data were available were broadly representative of the larger SEEM cohort, as shown in **S Table 1**.

**Figure 1:**
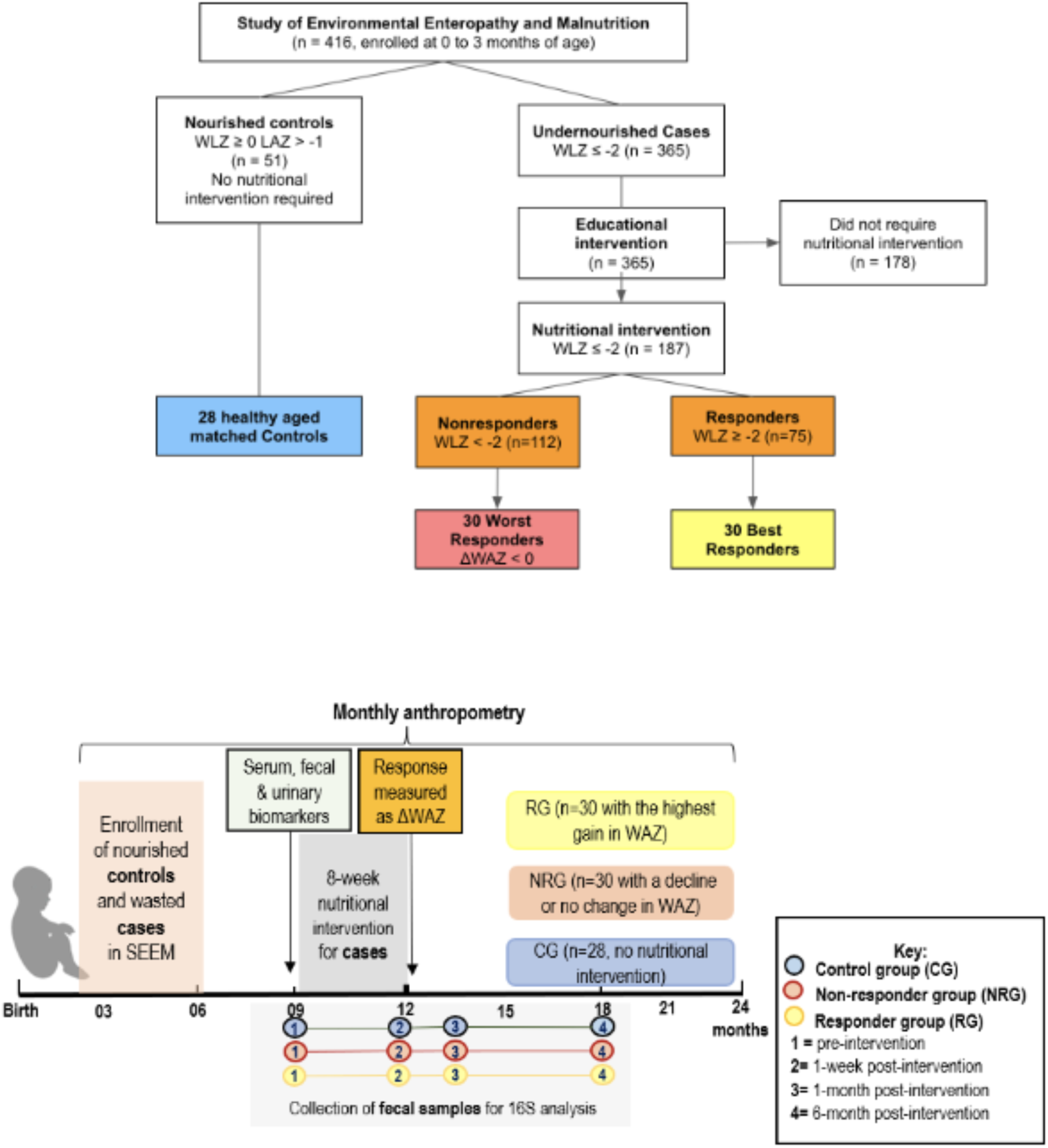
Overview of study design. **A:** Flow diagram of study population and protocols **B:** Infogram of study timelines and sample collection.

**Table 1:**
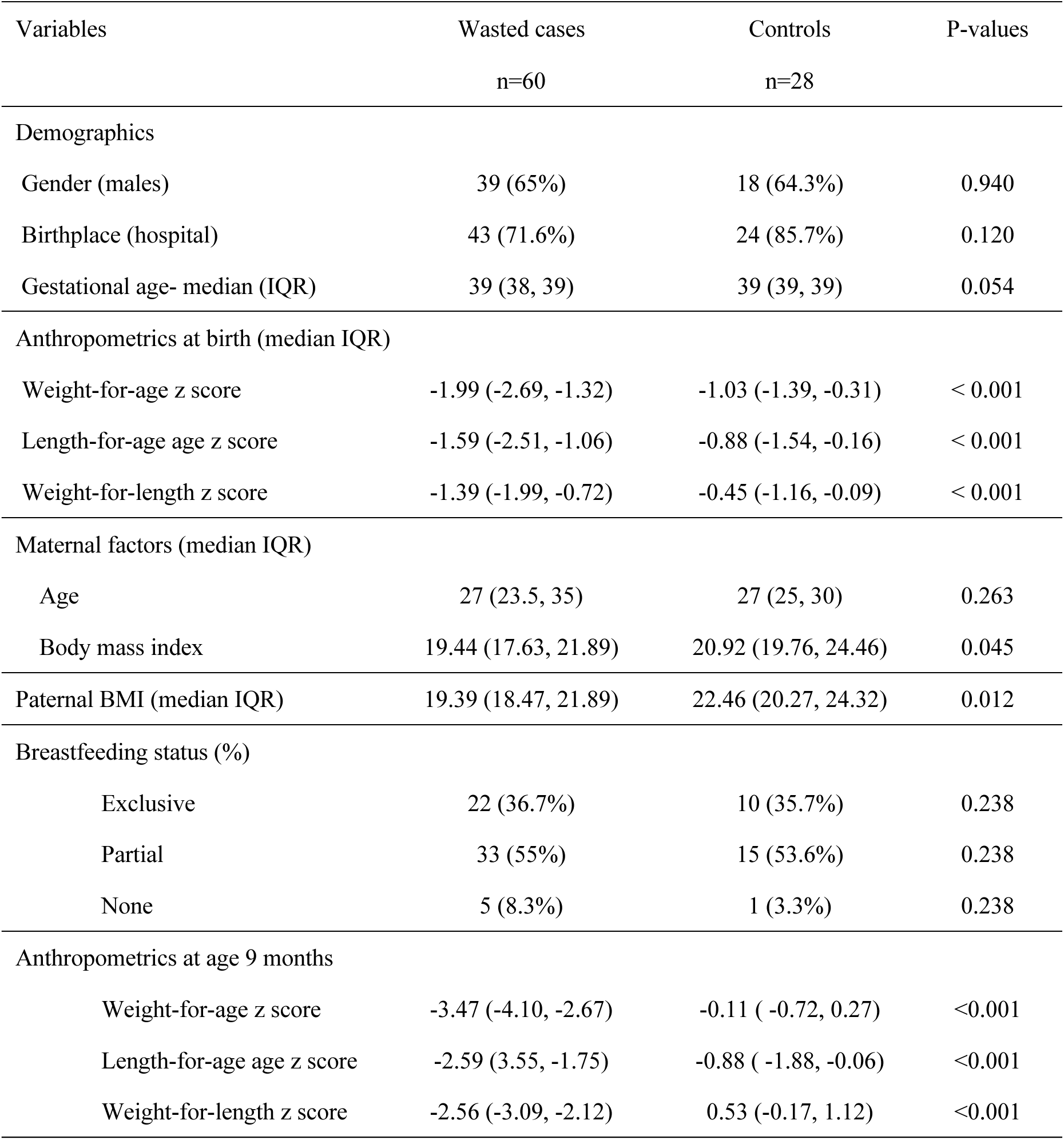
Baseline characteristics of Study participants (n=88)

Beginning at age 9 months, wasted infants (WLZ < -2, n = 187) whose growth did not improve after their primary caregiver participated in an intensive nutritional education intervention were provided Acha Mum, a locally produced chickpea-based RUSF, daily for 8 weeks. We collected blood, fecal, and urine samples prior to initiation of RUSF, allowing us to determine baseline EED biomarker and 16S fecal microbiome profiles (**Fig 1B**). We performed 16S sequencing of fecal specimens from four timepoints: (pre-intervention (age 9 months), one week post-intervention (age 12 months), one month post-intervention (age 13 months), and six months post-intervention (age 18 months) in three subgroups of participants: 1) the 30 best RUSF responders (RUSF-R), 2) the 30 worst RUSF non-responders (RUSF- NR), and 3) 28 controls who did not require nutritional interventions during the study. To quantify RUSF response, we calculated WAZ velocity (ΔWAZ/month) during the course of the 8-week intervention rather than WLZ velocity, given that changes in infant length are more challenging to reliably measure and detect over an 8-week nutritional intervention relative to changes in infant weight.

### Ponderal growth responses to RUSF administration

RUSF resolved wasting within one week of completing the 8-week intervention in only 41% of cases [10]. Of the 187 wasted infants who received RUSF, 75 (41%) showed improvements in WAZ (positive response); 112 (59%) did not. The 30 best responders (RUSF-R) demonstrated a mean WAZ velocity of 0.413 over the course of the intervention (**Fig 2A**, **Table 2**). RUSF-R showed significant improvements in overall nutritional status; severe acute malnutrition (SAM) declined from 33% to 0% and moderate acute malnutrition (MAM) declined from 63% to 20% (**Fig 2B**). In contrast, the 30 worst nonresponders (RUSF-NR) demonstrated either no improvements in WAZ or a decline, with a mean monthly WAZ velocity over the intervention of -0.081. The proportion of children with SAM increased in the RUSF-NR from 27% at baseline to 57% post-intervention. WAZ and WLZ scores were slightly lower in RUSF-R at baseline, indicating RUSF response was neither due to better nutritional status at baseline nor regression to the mean. Of note, RUSF-R were less wasted at birth relative to RUSF-NR (**S Table 1**). All other baseline indices were similar between groups, including RUSF adherence and illness burden ( **Fig S1A**).

**Figure 2:**
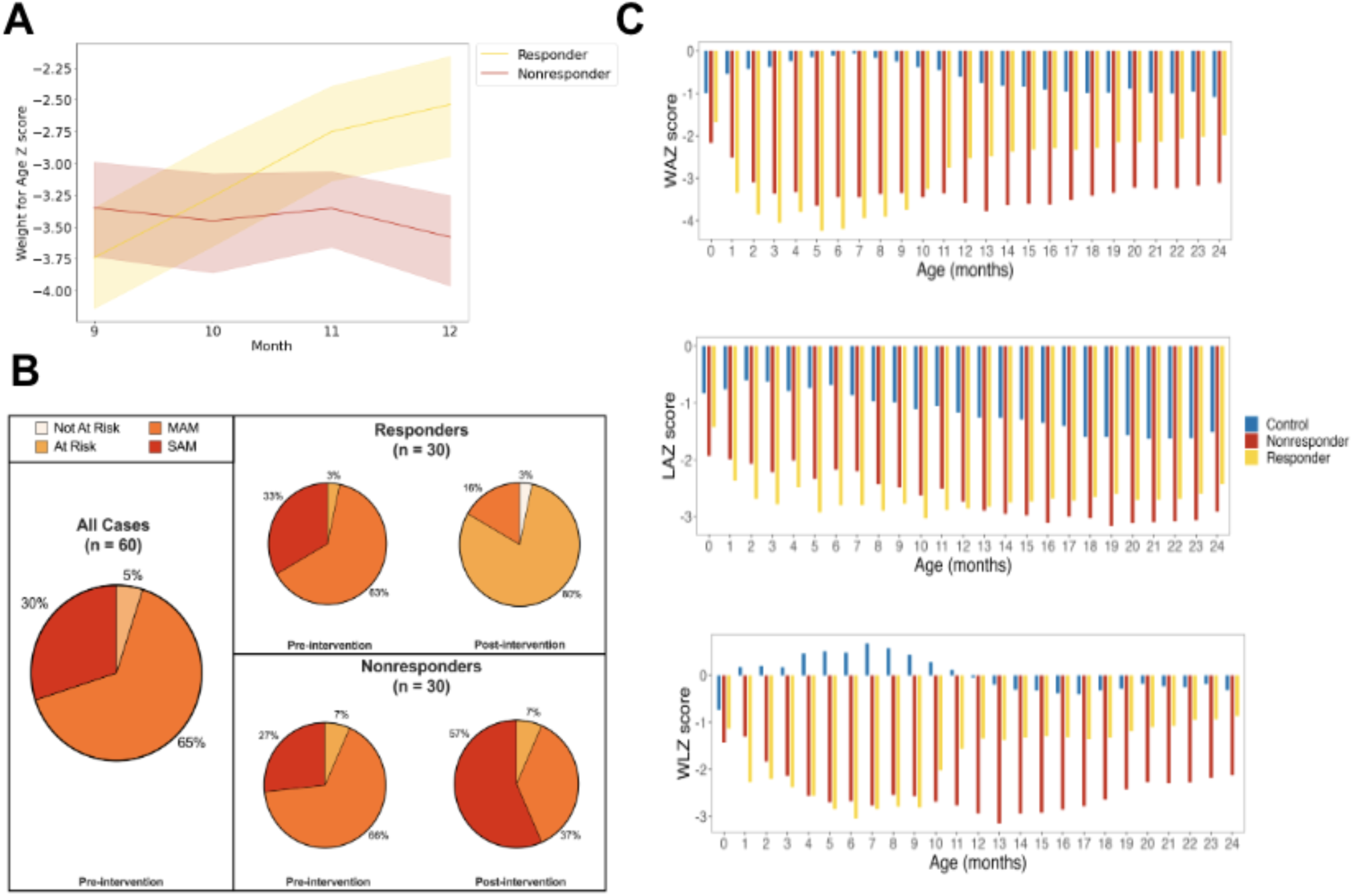
Children selected for gut microbiome analysis show diverging yet durable responses to RUSF. **(A)** Mean change in WAZ score per month during RUSF. **(B)** Nutritional status of responders and nonresponders with 16S data before and after RUSF. **(C)** On average, responders had worse ponderal growth prior to RUSF but showed persistent growth improvements in response to RUSF as measured by weight-for-age Z scores, length-for-age Z scores and weight-for-length Z scores. Age and region-matched well-nourished control cohort shown in blue.

**Table 2:** Clinical response to the RUSF supplementation in responders and nonresponders.

| Anthropometric features | Responders (n=30) | Nonresponders (n=30) | Difference (95% CI) † |
| --- | --- | --- | --- |
| **At baseline (pre-intervention) (mean ± SD)** |  |  |  |
| Weight-for-age z score | -3.740 ± 1.140 | -3.348 ± 1.075 | -0.392 (-0.989, 0.204) |
| Length-for-age z score | -2.774 ± 1.393 | -2.487 ± 1.260 | -0.287 (-1.002, 0.428) |
| Weight-for-length z score | -2.803 ± 0.702 | -2.564 ± 0.858 | -0.239 (-0.661, 0.183) |
| **Mean rate of growth per month from baseline to end of intervention (95% CI) †** |  |  |  |
| Weight-for-age z score | 0.413 (0.360, 0.466) | -0.081 (-0.133, -0.028) | 0.494 (0.401, 0.588) |
| Length-for-age z score | -0.006 (-0.048, 0.037) | -0.082 (-0.124, -0.039) | 0.076 (-0.005, 0.157) |
| Weight-for-length z score | 0.481 (0.398, 0.563) | -0.130 (-0.212, -0.048) | 0.611 (0.500, 0.722) |
| **Mean rate of growth from baseline to six months of follow-up (95% CI) ‡** |  |  |  |
| Weight-for-age z score | 0.046 (0.011, 0.081) | 0.022 (-0.013, 0.058) | 0.024 (-0.035, 0.083) |
| Length-for-age z score | 0.033 (-0.001, 0.066) | -0.055 (-0.085, -0.026) | 0.088 (0.044, 0.133) |
| Weight-for-length z score | 0.009 (-0.041, 0.058) | 0.053 (0.008, 0.097) | -0.044 (-0.111, 0.023) |

†Values for between-group difference at baseline were derived from a linear model predicting anthropometric features at start of treatment as a function of treatment group. Values for between-group difference in growth rate per month during the treatment period and during follow-up were derived from a mixed-effects linear model predicting anthropometric features as a function of the interaction between treatment group and the number of weeks since the initiation of nutritional supplementation after adjustment for the baseline variables as well as number of weeks of treatment, treatment group, and a random intercept for each participant to account for the within-participant correlation. Positive values indicate a faster growth rate in responders.
‡ Values for the rate of growth per month during six months of follow-up were derived from a mixed-effects linear model. This model predicts anthropometric features as a function of the number of weeks since the initiation of nutritional supplementation after adjustment for the baseline variables plus a random intercept for each participant to account for the within-participant correlation.

We tracked growth trajectories in RUSF-R and RUSF-NR beyond completion of supplementation (**Fig 2C**). Improvements in WAZ and WLZ continued in RUSF-R after the intervention was completed, persisting until the end of follow-up at 24 months, indicating a durable RUSF effect. RUSF-NR did not respond immediately to nutritional intervention; however, we noted improvements in ponderal growth beginning at age 13 months (**Table 2**). Length-for-age Z (LAZ) scores remained largely unchanged in both RUSF-R and RUSF-NR over the course of intervention and during follow-up. In controls who did not require nutritional intervention, anthropometric indices gradually declined over the course of follow-up. Taken together, this suggests that RUSF was ultimately beneficial even for RUSF-NR.

### Serum, urine, and fecal biomarkers distinguish wasted children from controls

We hypothesized baseline biomarkers of growth, inflammation, and EED would be distinct in wasted infants compared to well-nourished controls. All available biomarker data from the entire cohort (n = 235) were compared using Principal Component Analysis (PCA) to identify global group differences (**Fig 3A**). Our model was able to separate cases and controls along Principal Component 2 (PC2), with variation driven by biomarkers of growth and nutritional status (IGF-1, prealbumin, GLP-2, and leptin) as well as acute phase proteins C-reactive protein (CRP) and AGP (**Fig S2A**). Control samples clustered tightly, whereas cases displayed a more heterogeneous distribution along PC1, with a subset of participants designated as cases regardless of nutritional status. PC1 represented most of the variation within the model, primarily driven by inflammatory cytokines such as IL-1b, IFN-γ, and TNF-α. The PCA of the 88 study participants, selected for RUSF sub analysis (30 RUSF-R, 30 RUSF-NR, and 28 controls), showed similar a pattern of clustering (**Fig S2B)**, indicating that biomarker profiles from these subgroups were broadly representative of the total cohort.

**Figure 3:**
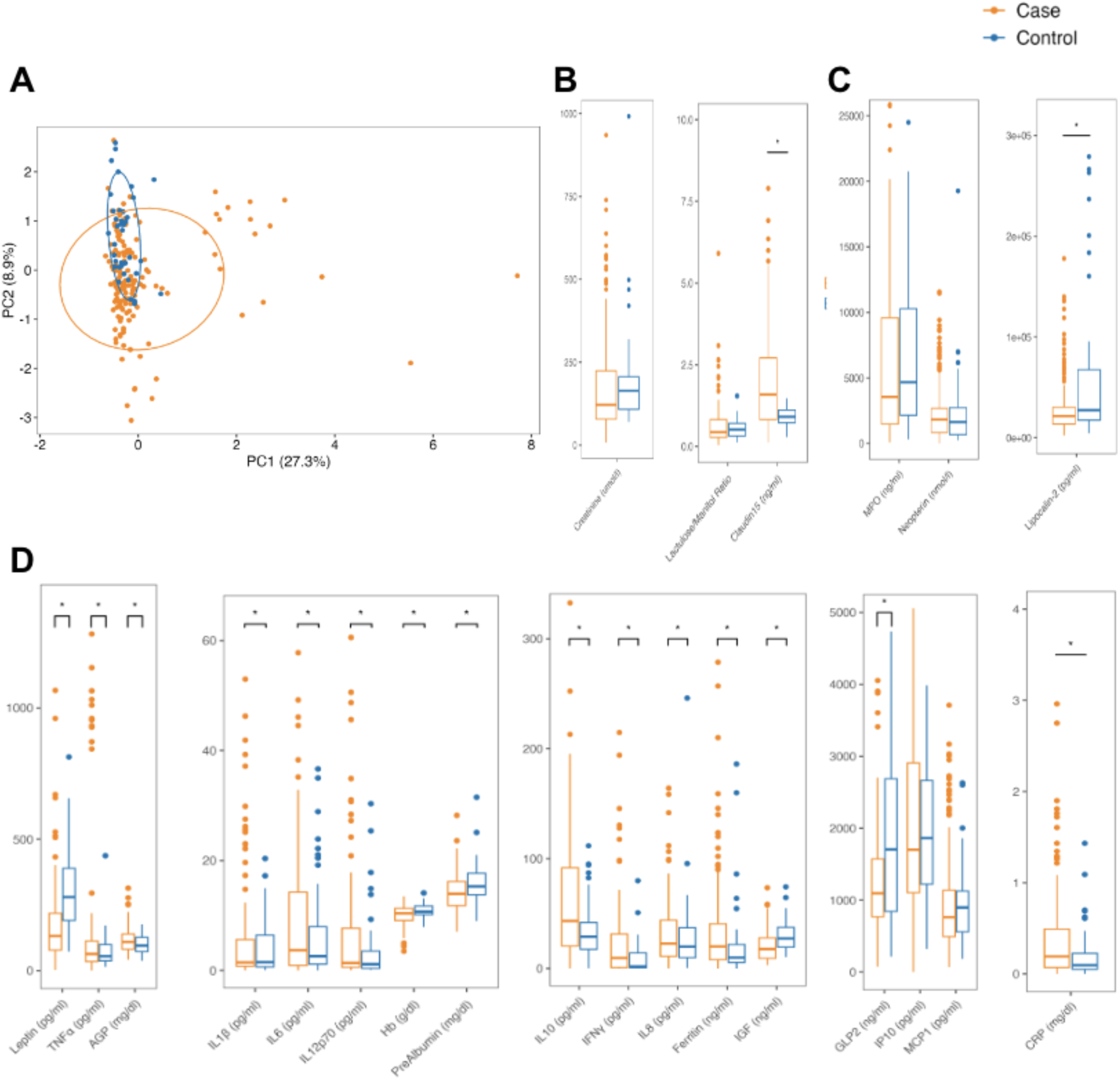
Stunted children exhibit increased systemic inflammation compared to healthy age-matched controls. **(A)** Principal component analysis of biomarker profiles from stunted children (n=148) and healthy aged-matched children (n=39). **(B)** Urine, **(C)** fecal, and **(D)** serum biomarker/cytokine levels from children at 9 months of age prior to RUSF in the SEEM cohort.

PCA findings were confirmed by univariate analyses. Biomarkers of growth and nutritional status, including IGF-1, prealbumin, GLP-2, and leptin, were significantly elevated in control samples compared to cases (**Fig 3B-D**). Cases displayed evidence of systemic inflammation, with significantly higher concentrations of inflammatory biomarkers such as CRP, IL-6, IFN-γ, TNF-α, and IL-1β. Of note, the anti-inflammatory cytokine IL-10 was also higher in cases vs. controls, suggesting feedback regulation. In addition, the urinary excretion of the intestinal tight junction protein claudin-15 was significantly higher in cases, consistent with previous associations of urinary claudin-15 with decreased gut barrier function observed in EED [11, 12]. Fecal markers of increased intestinal inflammation were similar across cases and controls: fecal myeloperoxidase (MPO) and neopterin (NEO) levels were similar between groups, and fecal lipocalin-2 (LCN2) was elevated in controls compared to cases.

### Pre-Intervention Biomarkers Predict RUSF Response

The heterogeneity of biomarker profiles observed in cases suggested a mixed population among this group, therefore we hypothesized biomarkers would help identify which undernourished infants were less likely to respond to RUSF. Relabeling cases by nutritional response (RUSF-R vs. RUSF-NR) supported this hypothesis; the majority of samples that failed to cluster with control samples belonged to the nonresponder group (**Fig 4A**). Heterogeneous samples were more likely to be RUSF-NR; however, RUSF-R and RUSF-NR groups as a whole did not cleanly separate, indicating overall broad similarity in biomarker profiles. In univariate analyses, fecal NEO, serum CRP and IL-10 were significantly higher in RUSF-NR vs. RUSF-R prior to intervention, however, we identified fewer significant differences than what we observed between cases and controls (**Fig 4B-D)**. Therefore, we took a machine learning approach to identify biomarkers more predictive of RUSF response.

**Figure 4:**
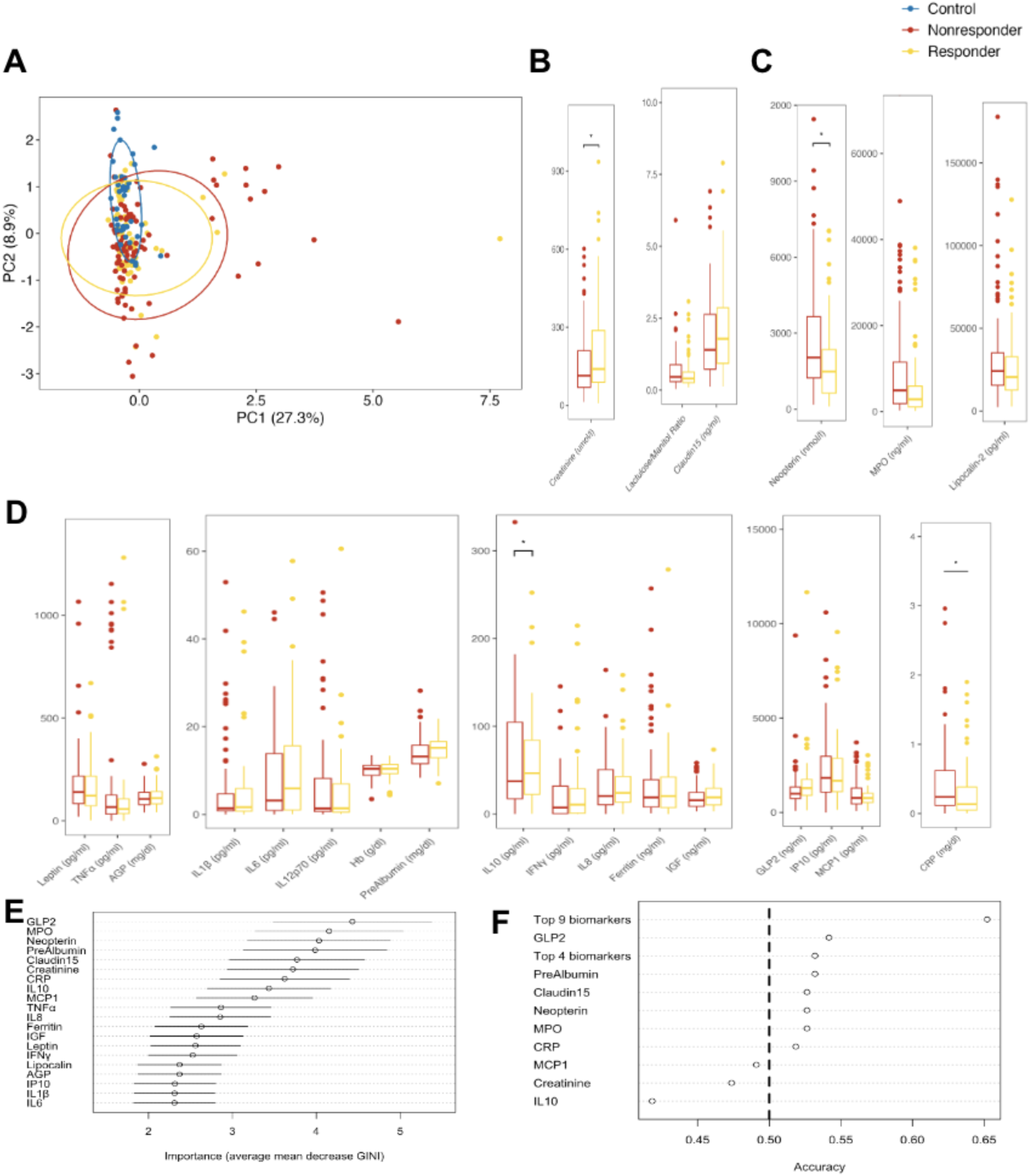
Baseline inflammatory biomarkers predict response to intervention: **(A):** Principal component analysis displaying the relationship between biomarkers and cytokines in the responders (n=66), nonresponders (n=82) and controls (n=38) at 9 months of age. **(B)** Urine, **(C)** fecal, and **(D)** serum biomarker levels from stunted children at 9 months of age prior to RUSF in SEEM cohort. Asterisk indicating p < 0.05. **(E)** Variable importance as shown by mean average decrease in GINI for each biomarker in Random Forest Model trained to predict whether a child would respond to the nutritional intervention. Circles indicate average importance; bars indicate standard deviation of 20-fold cross validation. **(F)** Accuracy of logistic regression models trained to predict response from individual biomarkers or combinations of biomarkers deemed important from Random Forest analysis.

Biomarkers were incorporated into an ensemble of Random Forest (RF) models with 20-fold cross-validation. Using this approach, biomarker data was able to correctly predict response to nutritional intervention with 60% accuracy (**Fig 4E**). A distinct set of nine features (GLP-2, MPO, prealbumin, NEO, claudin-15, creatinine, CRP, IL-10, and MCP-1) representing a mix of growth and inflammatory biomarkers were identified as most important for distinguishing responders from nonresponders. We observed a trend towards increased markers of growth and nutritional status (GLP-2, prealbumin, creatinine) in responders, consistent with the elevation of these markers in control samples relative to cases. We also noted a trend towards increased intestinal inflammation (MPO, NEO) in nonresponders–a difference not seen in comparisons of cases and controls. IL-10 was significantly higher in responders and CRP was significantly higher in nonresponders, highlighting elevated systemic inflammation as predictive of failure to respond. To interrogate the importance of individual biomarkers, a logistic regression model was trained based on these nine biomarkers. This model correctly predicted response with 68% accuracy on training set data and 73% accuracy on withheld test set data (**Fig 4F)**. Regression models built using either individual biomarkers or the collective top 4 biomarkers according to random forest performed poorly, with accuracy ranging from 50-60%. Broadly, these results indicate that biomarkers of growth, in conjunction with biomarkers of intestinal and systemic inflammation, can reliably distinguish RUSF responders and nonresponders prior to intervention.

### Gut microbiome diversity differs between wasted children and controls

Next, we explored the fecal microbiome community profiles of cases (including responders and nonresponders) and controls. We performed 16S rRNA sequencing on stool samples collected at baseline (pre-intervention) and at several time points post-intervention to identify amplicon sequence variants (ASVs), representing unique biological sequences that can be assigned a taxonomy. We first looked broadly at group differences in alpha (within the sample) and beta (between samples) diversity. Alpha diversity was significantly lower in controls compared to cases (**Fig 5A**). This was true when measuring observed ASVs as a measure of richness (how many unique taxa are present) and when measuring Simpson’s Diversity, which accounts for both richness and evenness (how evenly the relative abundance of taxa are distributed). Cases and controls also displayed significantly different beta diversity, as visualized using nonparametric multidimensional scaling (NMDS) on a Bray-Curtis Dissimilarity matrix (**Fig 5B**). These results suggest that wasted children in our cohort had a distinct microbiome from controls, characterized by increased diversity and greater evenness of taxa.

**Figure 5:**
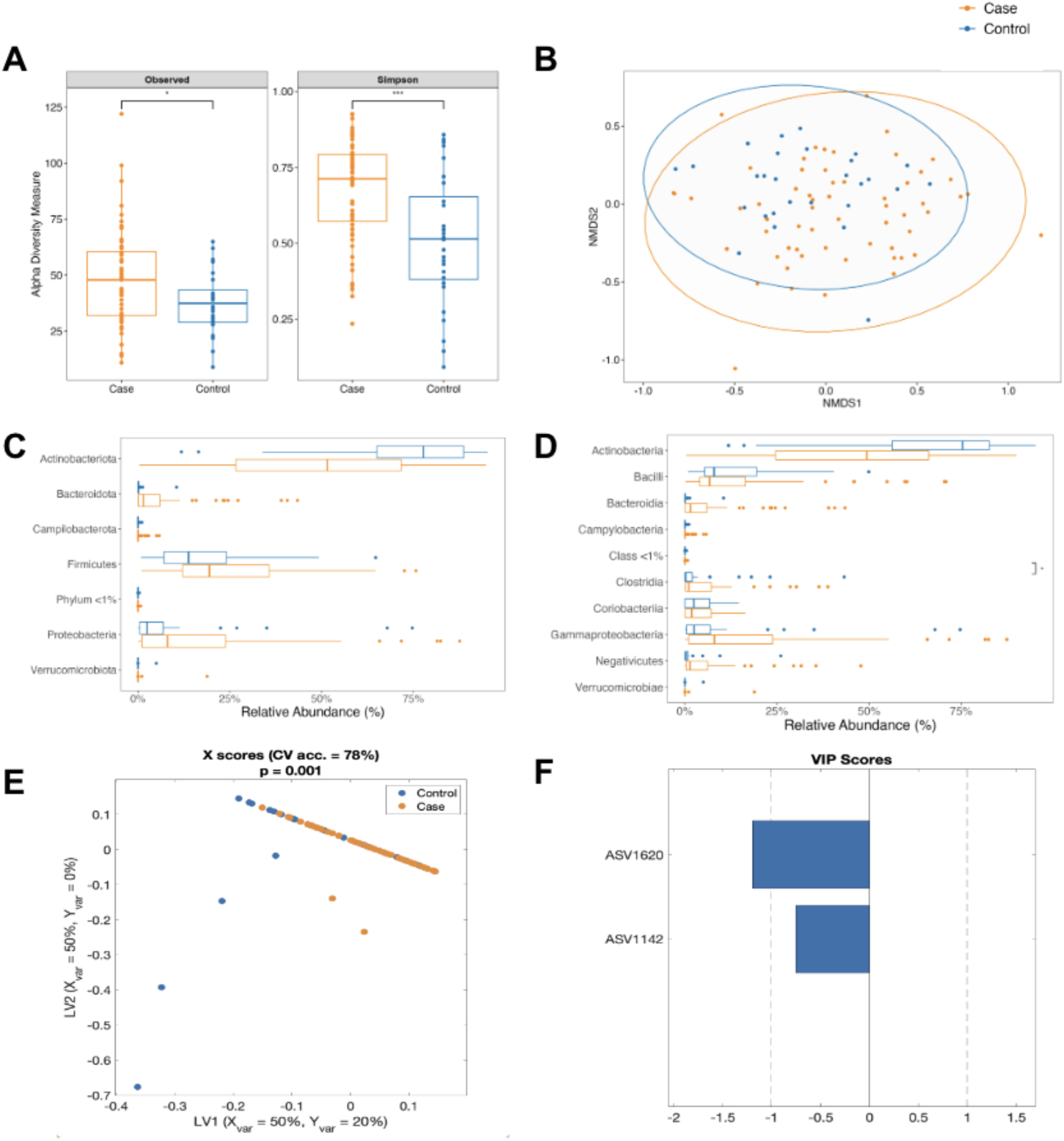
Wasted microbiota are more diverse relative to well-nourished microbiota: **(A)** Alpha diversity measurements (Number of observed species, Simpson index) of wasted cases and healthy controls at 9 months of age prior to RUSF. (**B):** Non-metric multidimensional scaling plot on Bray-Curtis matrices comparing the community composition of the cases and controls prior to the intervention. Relative abundance of **(C)** Phylum and **(D)** Class level taxonomy between cases and controls prior to the nutritional intervention. An OPLSDA model was constructed to discriminate between cases and controls using the relative abundance of ASVS from their fecal microbiome. The model outperformed all of 1000 randomly permuted models (p < 0.001). **(E)** Scatter plot of the X scores on latent variables 1 and 2 (LV1 & LV2), where each point represents one sample. **(F)** Bar plot shows the Variable Importance in Projection (VIP) scores, artificially oriented in the direction of loadings on LV1 and colored according to their association with case or control samples. VIP scores > 1 indicate a variable with greater than average influence on the projection.

We next determined which specific phylogenetic groups were different between cases and controls. Consistent with the decreased diversity observed in controls versus cases, the community composition of control samples was largely dominated by Actinobacteriota, whereas this phylum made up a significantly smaller proportion of the total community in cases (**Fig 5C**). The lower relative abundance of Actinobacteriota in cases was compensated by a significant increase in Proteobacteria and trends towards increased *Firmicutes* and Bacteroidota relative to controls. These changes were also reflected at the class level (**Fig 5D**), with a relative decrease in Actinobacteria in cases and increased *Gammaproteobacteria* and trends towards increased Bacteroidia, Negativicutes, and Clostridia.

Finally, a LASSO regularized, orthogonal Partial Least Squares-Discriminant Analysis (OPLS-DA) model was constructed to discriminate between case and control samples and identify ASVs that best discriminate between groups (**Fig 5E**). The model outperformed all 1000 randomly permuted models (p < 0.001) and achieved a cross-validation accuracy of 78% **(Fig S3A)**. Variable Importance in Projection (VIP) scores were plotted for top features and reflected ASV important for discriminating between groups (absolute value of VIP >1 is an important feature, with the sign of VIP indicating which group it is related to) **(Fig 5F)**. This analysis identified ASV1620, a member of the *Bifidobacterium* genus, as particularly important in discriminating between cases and controls. Univariate analysis showed ASV1620 was significantly enriched in control samples (**Fig S3B**). This ASV is a member of the Actinobacteriota phylum and *Bifidobacterium* genus, which are known to be important in early life [21]. As LASSO hides features that are linearly correlated to key driver features, ASVs with high correlation to those identified in the model may also be significant and are shown in a correlation network (**Fig S3C**). Taken together, our results show greater microbiome diversity at the expense of *Bifidobacteria* species is a signature feature of undernourished cases prior to RUSF.

### ASV-level differences reliably distinguish responders from non-responders among wasted children

We next sought to identify compositional differences in gut microbial communities of RUSF responders and nonresponders—both baseline differences and longitudinal changes post-intervention. We observed no significant differences in either alpha or beta diversity between responder and nonresponders prior to RUSF **(Fig 6A-B**), indicating the gut microbiota of these groups are broadly more similar to one another, relative to more striking diversity differences between cases and controls. Community composition in RUSF-NR mostly consisted of enrichment in Actinobacteriota, whereas RUSF-R exhibited a significantly lower proportion of Actinobacteriota and increased Proteobacteria (**Fig 6C-D**), similar to the differences we observed in cases versus controls. Parallel trends were seen at the class level, where the relative abundance of Actinobacteria and Gammaproteobacteria were significantly higher and lower in RUSF-R. In addition, we noted trends towards a higher relative baseline abundance of Firmicutes at the phylum level and Negativicutes at the class level.

**Figure 6:**
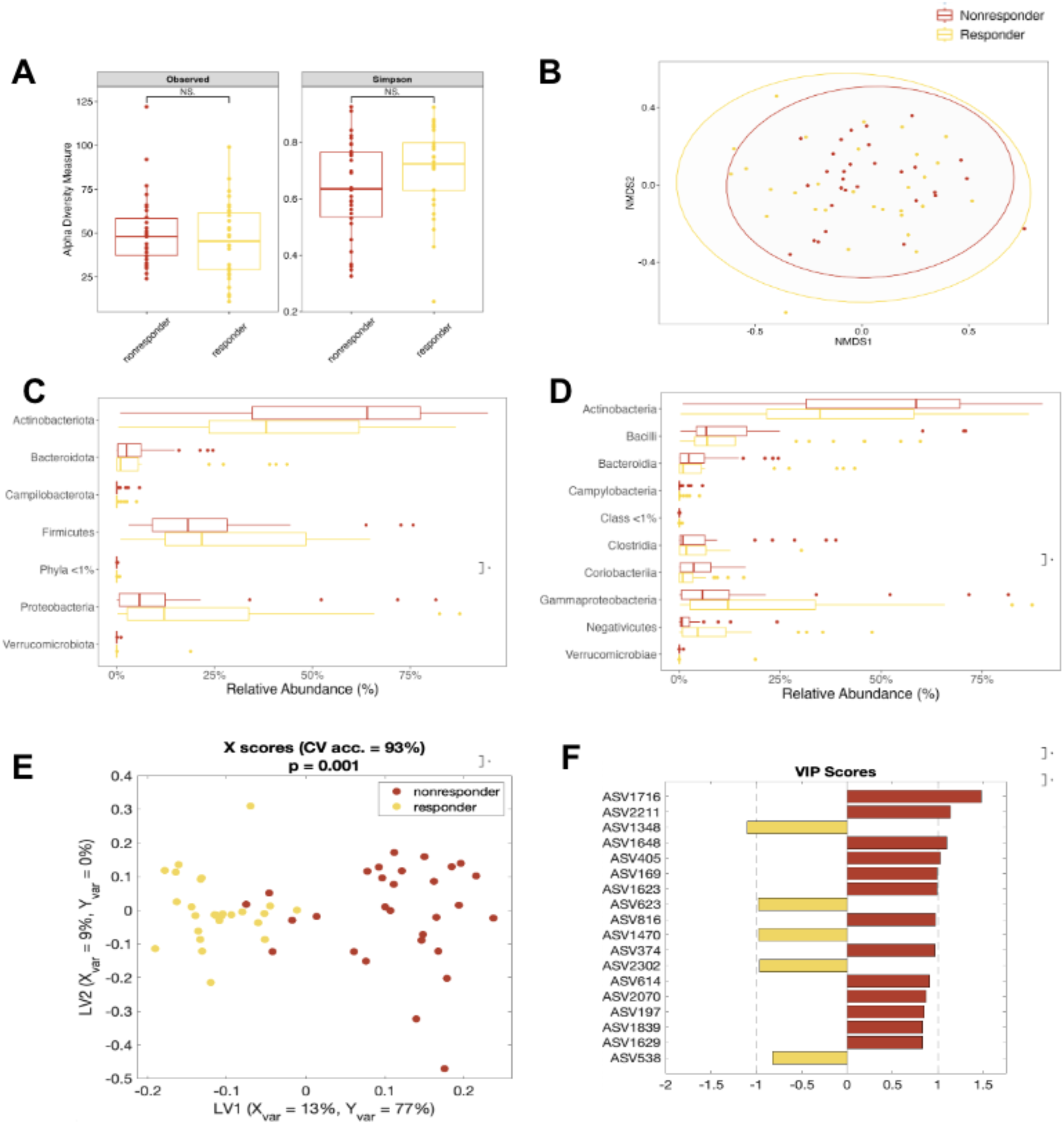
Responders and nonresponders harbor distinct fecal microbiota prior to RUSF. **(A)** Alpha diversity measurements (Number of observed species, Simpson index) of responders and nonresponders at 9 months of age prior to RUSF. (**B):** Non-metric multidimensional scaling plot on Bray-Curtis matrices comparing the community composition of the responders and nonresponders prior to the intervention. Relative abundance of **(C)** Phylum and **(D)** Class level taxonomy between cases and controls prior to the nutritional intervention. An OPLSDA model was constructed to discriminate between responders and nonresponders using the relative abundance of ASVS from their fecal microbiome. The model outperformed all of 1000 randomly permuted models (p < 0.001). **(E)** Scatter plot of the X scores on latent variables 1 and 2 (LV1 & LV2), where each point represents one sample. **(F)** Bar plot shows the Variable Importance in Projection (VIP) scores, artificially oriented in the direction of loadings on LV1 and colored according to their association with responder or nonresponder samples. VIP scores > 1 indicate a variable with greater than average influence on the projection.

We then generated a LASSO regularized, orthogonal Partial Least Squares-Discriminant Analysis (OPLS-DA) model to distinguish responders from nonresponders based on gut microbiome composition prior to RUSF. Remarkably, this model outperformed all 1000 randomly permuted models (p < 0.001) and achieved a cross-validation accuracy of 93%, indicating distinct differences at the ASV level between RUSF-R and RUSF-NR (**Fig S4A)**. VIP scores were plotted and used to determine ASVs important for discriminating between groups (**Fig 6E-F)**, with univariate analysis showing significant enrichment in several of the important features (**Fig S4B).** ASVs with a high correlation (70%) to those identified in the model that may also be of significance are shown in a heatmap (**Fig S4C**). This analysis points to a distinct pre-RUSF microbiome state characterized by increased colonization of Proteobacteria in children whose ponderal growth markedly improved after RUSF.

### Responders display shifts in gut microbiome composition during RUSF; microbiome composition in responders and nonresponders converge in the aftermath of RUSF

We next turned our attention to longitudinal changes in the microbiome over RUSF supplementation. Alpha diversity increased during the intervention among responders and nonresponders (**Fig 7A**), a pattern also observed in controls (**Fig S5A**). We identified no significant shifts in RUSF-NR microbiota at the phylum or class levels. In contrast, in RUSF-R, we identified a significant decline in the relative abundance of Proteobacteria, largely driven by reductions in Gammaproteobacteria and an increase in Clostridia. Importantly, we observed a robust increase in the relative abundance of Negativicutes in RUSF-R relative to RUSF-NR (**Fig S5B**) pre- and post-RUSF (**Fig 7B, Fig S7C**). In RUSF-NR, taxa were not immediately different pre- vs. post-intervention. At the family level, shifts in the Negativicutes class were driven by members of *Veillonellaceae*, Proteobacteria by members of *Enterobacteriaceae,* and Actinobacteria by members of the *Bifidobacteriaceae* family (**Fig** 7**C**).

**Figure 7:**
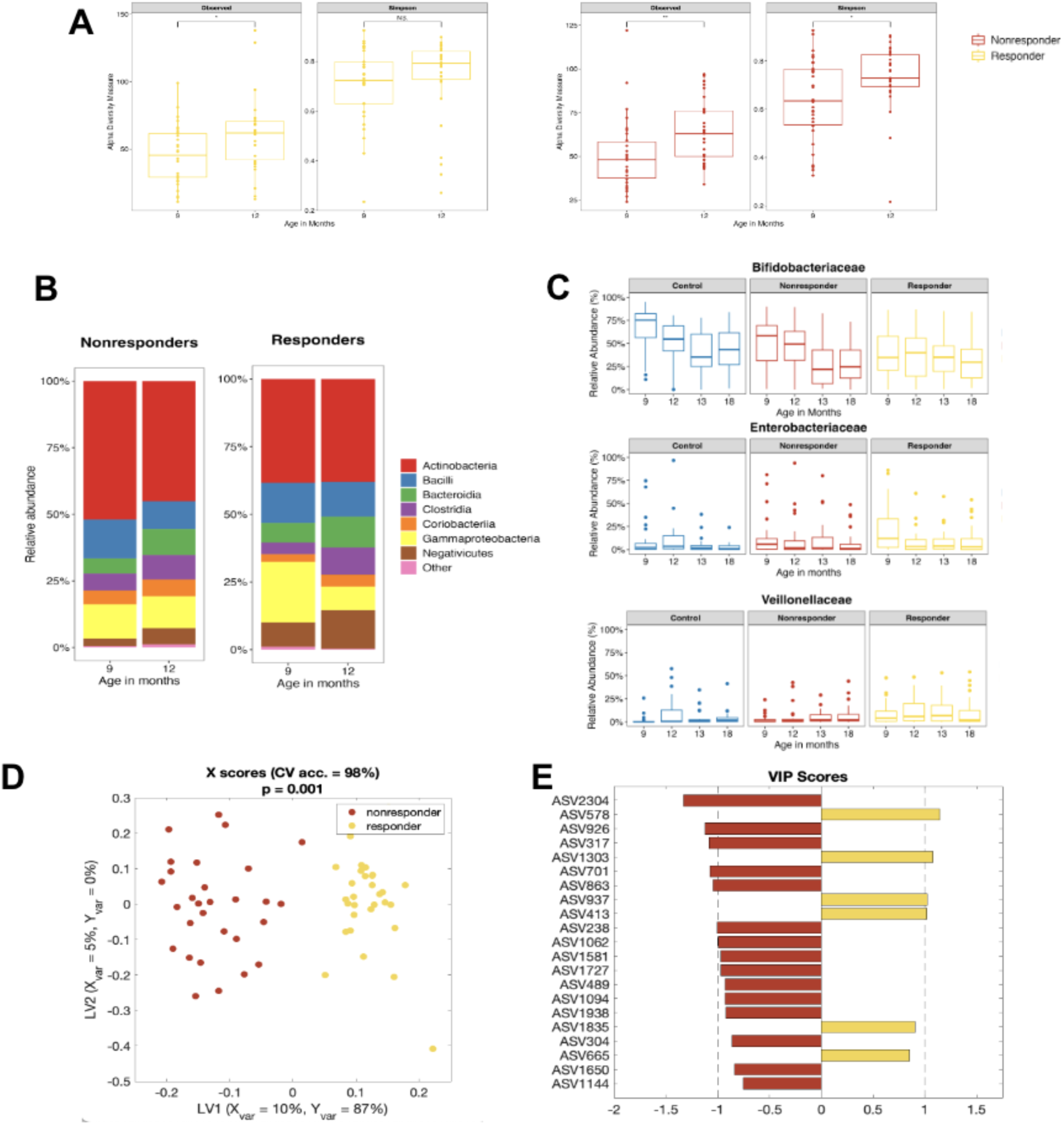
RUSF responders exhibit distinct shifts in gut microbiome composition over the course of the nutritional intervention. **(A)** Alpha diversity measurements (Number of observed species, Simpson index) of responders and nonresponders pre and post intervention. **(B)** Comparison of relative abundance of microbial taxa at the phylum level in responder and nonresponder stool samples over the course of the study. **(C)** Comparison of relative abundance of select microbial taxa at the family level in responder, nonresponder, and control stool samples throughout the course of the study. An OPLSDA model was constructed to discriminate between responders and nonresponders using the relative abundance of ASVs from their fecal microbiome. The model outperformed all of 1000 randomly permuted models (p < 0.001). **(D)** Scatter plot of the X scores on latent variables 1 and 2 (LV1 & LV2), where each point represents one sample. **(E)** Bar plot shows the Variable Importance in Projection (VIP) scores, artificially oriented in the direction of loadings on LV1 and colored according to their association with responder or nonresponder samples. VIP scores > 1 indicates a variable with greater than average influence on the projection.

To identify specific ASVs associated with RUSF response, we constructed a LASSO regularized OPLS-DA model to discriminate between RUSF-R and RUSF-NR samples immediately post-intervention (12 months of age) **(Fig 7D-E)**. The model outperformed all of 1000 randomly permuted models (p < 0.001) and achieved a cross-validation accuracy of 98% **(Fig S6A)**. A plot of VIP scores showed several ASVs as important in discriminating between responder and nonresponder samples post-intervention. ASV2304, a member of the *Lachnoclostridium* genus, was the most important feature of RUSF-NR samples **(Fig 7E)**. Univariate analysis showed this ASV to be present significantly more often in nonresponder samples than in responder samples. The most important microbial feature relevant to RUSF-R samples was ASV578, a member of the *Veillonella* genus and Negativicutes class that was present more abundantly in responders (**Fig S6B**). ASVs with a high correlation (>70%) to those identified in the model that may also be of significance are shown in a heatmap (**Fig S6C**). Together, this analysis shows a RUSF-driven transformation of the gut microbiome in RUSF-R distinct from that observed in RUSF-NR over the course of the nutritional intervention. This transformation is characterized by a robust decrease in Gammaproteobacteria and an increase in Negativicutes.

### Systemic inflammation identifies RUSF-NR outliers with responder-like microbiome profiles

Among the 30 worst RUSF-NR, we identified seven individual nonresponders with distinct biomarker profiles relative to either controls or RUSF-R. These profiles were primarily driven by a significant elevation in biomarkers of systemic inflammation (**Fig 8A**). We next asked whether these individuals harbored distinct microbiota relative to other RUSF-NR. The microbiomes of inflamed RUSF-NR showed greater similarity to RUSF-R than to non-inflamed RUSF-NR (**Fig 8B**). This was most evidenced by lower levels of Actinobacteriota and increased levels of Proteobacteria in inflamed versus non-inflamed RUSF-NR. Inflamed RUSF-NR also exhibited distinct growth trajectories, with higher WAZ scores prior to RUSF followed by marked decreases in WAZ during the intervention. Follow-up analysis also showed that these infants experienced higher rates of diarrheal episodes and acute respiratory infections (**Fig S7A-B, Supp Table 3**). These findings suggest that microbiome structure and function play an important role in RUSF response; however, systemic inflammation and acute illnesses can undermine that response.

**Figure 8:**
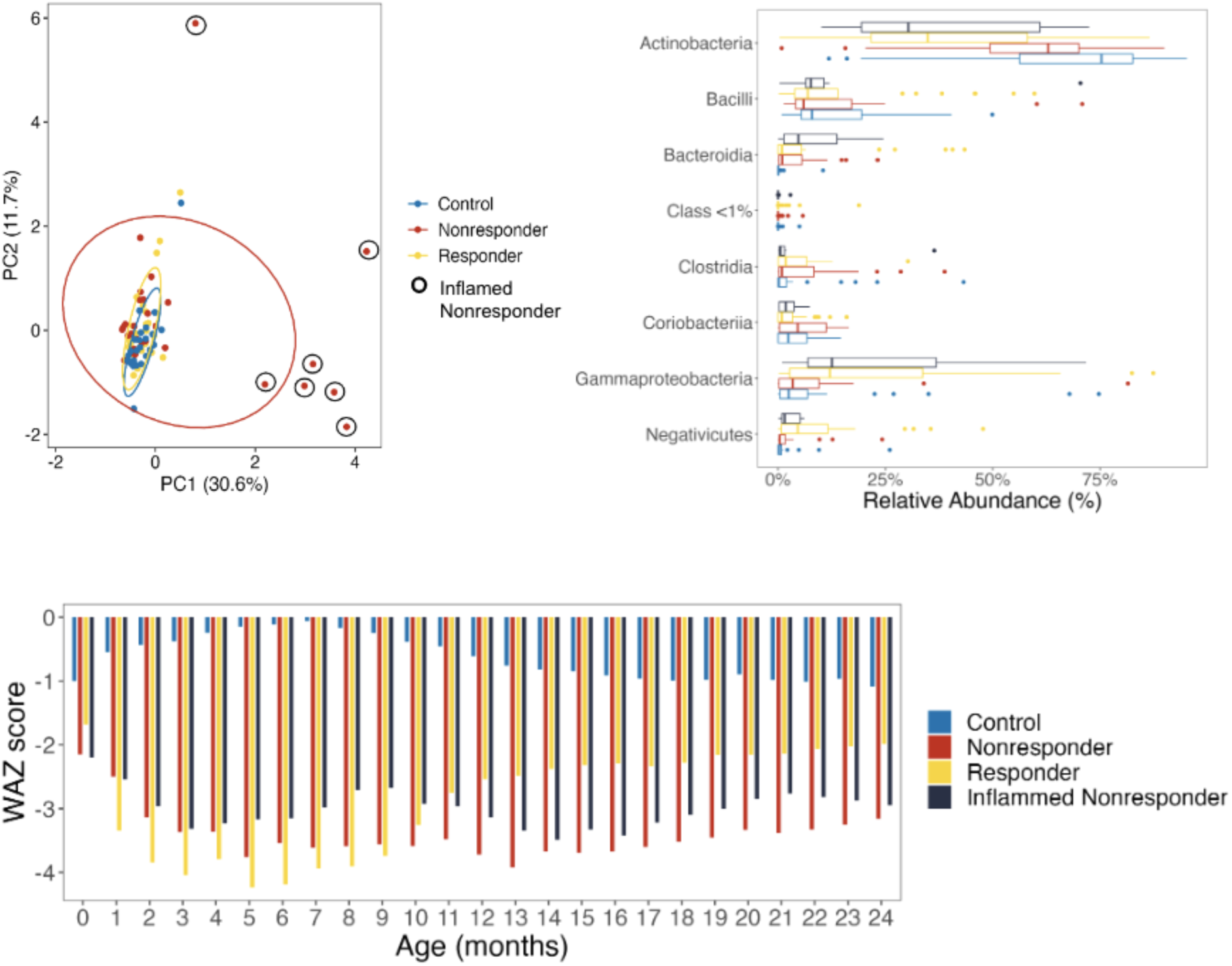
RUSF nonresponders with high levels of inflammation harbor fecal microbiota that resemble those of responders. **(A)** Principal component analysis displaying the relationship between biomarkers and cytokines measured pre-intervention from patients with matching microbiome data in the responders (n=30), nonresponders (n=30) and control groups (n=28). Black circles denote nonresponder samples with high levels of inflammation selected for downstream analysis. (**B)** Comparison of the average relative abundance of microbial taxa at the phylum level in responder, nonresponder, and inflamed nonresponder stool samples prior to the nutritional intervention. **(C)** Longitudinal change in WAZ score for control, responder, nonresponder, and inflamed nonresponder children over the first two years of life.

### Validation of the predictive model

After pinpointing microbial taxa with strong predictive potential, we validated our findings in an independent Bangladeshi cohort of young children. As limited studies were suitable for validation, we selected the 2022 study by Chen et al [13] due to its utilization of a chickpea-based MDCF. We utilized the data of 60 children who received the MDCF, and categorized them into three groups based on changes in WAZ during the intervention (**Fig S8A**). The fecal microbiome was compared between children showing the most favorable response and those showing the least. Unlike our dataset, at the pre-intervention stage, significant differences in Proteobacteria or Negativicutes abundance in the Chen et al. study were not seen (**Fig S8B)**. To uncover any consistent microbial taxa with predictive potential across cohorts, we first used principal component analysis on pre-intervention composition data (**Fig S8C**) and, seeing no significant batch effects, trained an OPLSDA model using pre-intervention microbiome data from the SEEM study and applied it to the pre-intervention validation dataset (**Fig S8D**). Although our model identified several taxa significantly linked to response, it only classified patients with 54% accuracy in the validation dataset. While this accuracy is only marginally better than chance, it underscores the microbiome composition variations between cohorts and studies.

## Discussion

We identified host-microbiome determinants of RUSF efficacy among a birth cohort of rural Pakistani infants at high risk for moderate and severe acute malnutrition. Through a comprehensive assessment of 1) baseline biomarkers of EED and systemic inflammation, 2) the structure and flux of gut microbial communities before and after RUSF, and 3) ponderal growth responses to RUSF, we uncovered several findings with important implications for the evaluation and treatment of acute childhood undernutrition in LMIC community settings. First, pre-RUSF biomarker profiles reliably distinguished RUSF responders from nonresponders; children with heightened gut and systemic inflammation were less likely to respond to supplementation. Second, we identified key baseline differences in the gut microbial community structure of nourished infants, RUSF responders, and RUSF nonresponders. Third, in contrast to nonresponders, RUSF responders displayed significant shifts in gut microbiota over the course of supplementation. Fourth, in a small group of outliers whose gut microbiome would have otherwise predicted positive RUSF response, high burdens of inflammation and infections undermined RUSF efficacy. Lastly, the predictive power of these and other measurable host-microbiome factors sets the stage for precision nutrition approaches to not only tailor the formulation of RUSF, or microbiome-directed complementary foods, to achieve greater efficacy but also shape the timing, duration, and adjunctive therapies to RUSF to promote optimal and durable growth outcomes.

RUSF responders weighed more at birth, further supporting the role of antenatal care and birth dynamics in establishing childhood growth trajectories [14]. Univariate comparisons of biomarkers revealed increased intestinal inflammation in RUSF-NR and increased markers of muscle mass (urinary creatinine) and the anti-inflammatory cytokine IL-10 in responders. Our models identified a set of nine biomarkers with acceptable predictive accuracy for children at risk of RUSF nonresponse. These included established biomarkers for growth (prealbumin, creatinine, leptin, GLP-2, and IGF-1) as well as commonly measured markers of gut and systemic inflammation (fecal MPO, serum AGP, and CRP). These biomarkers were tested because of the known microbial infections in environmental enteropathy [15, 16]. Together, increased growth biomarkers and anti-inflammatory cytokines may indicate that RUSF-R was primed to break out of the vicious cycle of inflammation and undernutrition once supplementation began. Our analysis provides valuable insights into the role of baseline biomarkers in identifying those who may require longer or additional therapeutic interventions in community settings.

RUSF responders and nonresponders displayed significant differences in microbiome composition pre-intervention. Although diversity indices were similar, responders harbored higher levels of Gammaproteobacteria and Negativicutes and lower levels of Actinobacteria prior to RUSF. A PLS-DA model trained to predict who would respond to RUSF based on the relative abundance of pre-intervention ASVs was able to classify responders with 93% accuracy, identifying several ASVs correlated with response versus nonresponse. Another key observation was that nonresponders exhibited minimal changes in gut microbiome composition, whereas responders displayed a high degree of microbiome restructuring over the course of supplementation. Gammaproteobacteria, a large class of bacteria that includes the pathobionts *Klebsiella*, *Salmonella*, and *Shigella*, were found in higher numbers before the intervention and decreased significantly thereafter in responders. We speculate that the high prevalence of these facultative anaerobes prior to intervention may have contributed to earlier growth faltering in these children. RUSF appeared to sufficiently restructure the microbiome of responders towards growth- conferring taxa, with significant increases in Negativicutes, including *Megasphera* and *Veillonella*.

Recent studies show these taxa ferment chickpeas to release beneficial metabolites [17], which might account for their increased presence in Acha Mum responders.

In concordance with prior observations that inflammation and repeated infections abrogate catch-up growth in childhood [18, 19], we found that nonresponders with the highest burden of inflammation, diarrhea, and respiratory infections harbored microbiomes more similar to responders rather than typical RUSF nonresponders. This may indicate that this small group of infants was primed to respond to RUSF but derailed by acute illness and inflammation. Hence, gut and systemic inflammation and the state of the gut microbiome may jointly coordinate the metabolic response to RUSF. Future studies should explore targeting the reduction of acute inflammation and/or infection before or during the administration of RUSF as a strategy to increase RUSF efficacy [20, 21].

We attempted to validate the findings of this study in another cohort of malnourished children. We found that while the pre-intervention time point data could predict response with high accuracy in this dataset, the key taxa that distinguished responders and nonresponders differed from our cohort. While the Chen et al dataset was the best match for our study and used a chickpea-based intervention, there were several key differences in their study design. First, while our study took place in an arid rural region in Pakistan, the validation study occurred in Dhaka’s humid urban environment. Second, while our study included children with both SAM and MAM, the validation study included only children with MAM. Finally, in our study, the intervention began at 9 months of life, whereas the validation study took place between the 12th and 18th months of life. Together, these factors may all contribute to the different trends observed in microbiome restructuring.

This study benefited from several strengths. First, we included well-nourished controls from the same rural settings who did not require RUSF, providing a set of local, age-matched controls. Second, we tested various blood, stool, and urine biomarkers to distinguish relative contributions from EED versus systemic inflammation. Third, the longitudinal collection of fecal samples enabled us to observe the restructuring of the microbiome and correlate this restructuring with the RUSF response. Lastly, by monitoring anthropometrics up to 24 months of age, we explored the intervention’s long-term effects and identified delayed onset improvements in ponderal growth among RUSF nonresponders. The limitations of our study include performing serial 16S rRNA fecal analyses on only 28 to 30 samples per group. Our focus on extreme RUSF responders and nonresponders to interrogate WAZ-associated microbial taxa may have missed more subtle host-microbiome interaction in children who showed a mild benefit from the intervention. We did not collect fecal samples during the intervention, which might have better captured the dynamics of microbiome transformation. With the limited sequencing depth provided by 16S rRNA analysis, confirming any potential probiotic/symbiotic candidates in gnotobiotic animal models or human nonclinical models, such as EED-on-a-chip [22], would be important prior to clinical interventions.

Lastly, although machine learning and other computational tools allowed us to identify important ASVs, the heterogeneous nature of gut microbiota means important features should be interpreted with caution.

For future studies of RUSF, we recommend exploring longer durations of intervention, especially in primary nonresponders—a group in whom we observed later convergence in the transformation of several ASVs. In responders, growth improvements with RUSF were durable; however, continued monitoring and support during critical growth periods is warranted. The correlations we identified between birthweight and RUSF response at 9 months of age underscore the importance of healthy pregnancy and birth anthropometrics in establishing growth trajectories. Our finding that common illnesses and inflammation undermined RUSF response re-emphasizes the importance of clean water, sanitation, and hygiene practices and access to essential drugs in optimizing the individual and community impact of RUSF. Adjunct therapy with probiotics tailored to RUSF or MDCF composition and the predominant infant gut microbial signatures of at-risk populations may enhance outcomes and is an ongoing area of study [23]. We suggest that future studies aimed at identifying bacteria associated with response to a nutritional intervention be designed to include multiple sites to better account for and understand how cohort setting impacts response. Finally, for children with either microbiome or inflammatory states associated with nonresponse, administering an antibiotic or anti-inflammatory medication prior to RUSF might conceivably prime the microbiome and gut for a more beneficial ponderal growth response.

We conclude that birth weight, gut and systemic inflammation, and gut microbial community composition profoundly influence the ponderal growth response of acutely undernourished infants to RUSF. The transformation of gut microbial community structure over RUSF supplementation further shapes this response. Multi-centered studies designed to validate promising biomarkers, microbial targets, and RUSF formulations–using harmonized methods across LMIC settings where the burden of acute and chronic childhood undernutrition is greatest–are urgently needed to develop scalable solutions toward global reductions in childhood wasting, underweight, and stunting.

## Material and methods

### Study participant details

Study participants were drawn from the SEEM-Pakistan birth cohort study (n=416) conducted in Matiari, Sindh. We selected participants for microbiome analysis based on the availability of longitudinally collected fecal samples. SEEM-Pakistan explored gut tissue histology, transcriptomics, and intestinal permeability in undernourished cases refractory to nutritional interventions.

The study was approved by the Aga Khan University Ethical Review Committee (ERC # 2021-0535- 19973), and written informed consent was obtained from the participant’s parents. The study was registered at ClinicalTrials.gov ID NCT03588013, and the SEEM protocols were published [24].

Newborns were enrolled at 0-3 months of age and followed monthly for growth monitoring. On the basis of two consecutive months of anthropometric measurements, 9-month-old infants were classified either as controls (WLZ ≥0, LAZ ≥-1) or wasted cases (WLZ ≤-2). Out of 350 cases, 187 were selected for RUSF nutritional intervention after not responding to a caregiver nutrition education program. Response to the Acha Mum RUSF intervention was monitored as improvement in WLZ ≥ -2 at one week after post-intervention, irrespective of their WLZ at the pre-intervention timepoint. 41% of cases responded in the SEEM study. For this study, of those 41% of cases who responded to the intervention, we selected 30 children with best response that is a maximum gain in WAZ (change > 0.5) and classified them as RUSF- R while from those 59% nonresponders; we selected the children (n=30) who experienced a decline in WAZ or no change and labeled them as RUSF-NR . The fecal samples of these thirty best responders and thirty worst non-responders based on the change in WAZ were selected for the 16S analysis in this sub-study.

### Data collection

Information on general demographic characteristics was collected by trained community health workers through interviews with the mothers, which included children’s birth data, breastfeeding history, and parental parameters. Anthropometric measurements were performed using standard procedure and equipment, with weight measured to the nearest 20g precision electronic scale (TANITA 1584) and length to the nearest 1 mm using a rigid length board with a movable footpiece. These measurements were collected monthly for up to 24 months and converted to z scores using the WHO Anthro 3.2 application. Based on established cut-offs for nutritional indicators, the participants were categorized as stunted (LAZ< -2), underweight (WAZ < -2), at risk of wasting (WLZ <-1 and ≥ -2), moderately wasted (WLZ < -2) and severely wasted (WLZ < -3), respectively.

### Sample collection for inflammatory biomarkers

Blood, fecal, and urine samples were collected from participants at nine months of age (**Fig. 1A**). 1 to 2 ml of blood was collected from which serum was aliquoted in small volumes to avoid freeze-thawing and transported at 4°C from the field site lab to the Infectious Disease Research Laboratory (IDRL) where they were stored at -80°C until processed.

Commercial ELISA kits were used for estimation of GLP-2 (USCN, Life Sciences Inc, Wuhan, China) while CRP, ferritin, and AGP were analyzed using a Hitachi 902 analyzer (Roche Diagnostics, Holliston, MA), and IGF-1 was measured using a LIAISON (Diasorin Saluggia (VC) Italy). All assays were performed following manufacturer protocols. For fecal samples, the child’s caretaker collected the sample using a wooden spatula and put it into a clean container provided in the stool collection kit. The fecal sample was transferred into multiple cryogenic vials for long-term storage. Commercial ELISA kits were used for the estimation of MPO (Immunodiagnostic AG, Stubenwald-Allee, and Bensheim) and NEO (GenWay Biotech, San Diego. CA). Fecal lipocalin (LCN2) was measured by DuoSet ELISA DY1757. All plates were read on Biorad iMark (Hercules, CA) plate reader.

For the evaluation of serum cytokines, a commercially available MILLIPLEX MAP Human Cytokine/Chemokine (MERCK) kit was used. The screening panel including IFN-γ (interferon- γ), IL-10, IL-12 (IL-12p70), IL-1β, IL-6, IL-8, IP-10 (Interferon-gamma-induced protein 10, also called as CXCL10), MCP-1(Monocyte chemoattractant protein-1, also called as CCL2) and TNF-α (Tumor necrosis factor) as per manufacturer’s instructions using Bioplex- 200 instrument. The data was analyzed using Bioplex Manager 6.1.

### Administration of nutritional intervention

Enrolled cases with WLZ < -2 at nine months were selected for nutritional intervention. Acha Mum, a ready-to-use supplementary food (RUSF), was given at a dose of one sachet per day to cases with MAM (WLZ scores between -2 and -3), while children with SAM (WLZ < -3) were administered sachets as per child’s weight (200 kcal/kg/day) [25]. Acha Mum composition is provided in the supplementary material. This eight-week intervention was monitored through weekly visits by the local team to document compliance, side effects, needs for medical assistance, and other details. Compliance was calculated (weekly) based on the empty wrappers returned by mothers [Compliance = (Total packet used/total packet given) *100]. One week after the completion of the intervention, the response was measured as the overall change in the WAZ of the children.

### Longitudinal fecal sample collection for 16S ribosomal RNA gene sequencing

Mothers of study participants were trained to collect the fecal samples and provided with mobile cards for timely communication with the collection team in the field. As described above, the mothers collected the fecal sample and sent an immediate message to the field team to pick up the sample within 30 minutes. Overall, four samples were collected from each child: one sample at nine months to capture microbiota at the pre-intervention stage, while three samples were collected at twelve, thirteen, and eighteen months to evaluate post-intervention taxa (**Fig. 1B)**. Fresh samples were transferred into a pre-chilled cryovial, snapped to the aluminum cryo cone and placed into a freezing container (Coleman) at 2° to 8°C. The time taken from the passage of fresh stools to being snap-frozen in a dry shipper was less than 30 minutes. The Coleman was carried in liquid nitrogen dry shipper to the local laboratory and later shifted to IDRL, AKU on dry ice where samples were stored at -80C until shipped to BGI Genomics (formerly Beijing Genomics Institute). A 30ng qualified DNA template was tested for sample integrity by agarose gel electrophoresis and concentration by a microplate reader (Qubit fluorometer). The quantified samples (6 -100ng/uL) were normalized to 30ng DNA per reaction. fusion primers were designed to include Illumina adapter sequences, an 8-nucleotide index sequence, and a gene-specific primer and added to the Polymerase chain reaction (PCR) reaction system. All PCR products were purified by Agencourt AMPure XP beads, dissolved in an Elution Buffer, and labeled for library construction. Library size and concentration were detected by Agilent 2100 Bioanalyzer. Qualified libraries were sequenced pair-end on the HiSeq 2500 platform according to their insert size. The raw reads were filtered to remove the adapter and low-quality bases. Paired-end reads were added to the tags by Fast length Adjustment of the Short read program (FLASH, v12.11). The sequence data are deposited in the NCBI Sequence Read Archive (SRA). Alpha rarefaction curves were generated to assess the effect of sampling depth on ASV abundance (S FigA). The plots indicate that the detection of ASVs had already attained a plateau of 60,000 reads. This trend was separately confirmed in the three groups.

### 16S ribosomal RNA gene analysis and statistical analysis

Sequence analysis was performed in R using DADA2 (version 1.22.0) [26]. The forward read was truncated to 200 base pairs, and reads with ambiguous ‘N’ bases and >2 expected errors were removed. Chimeras were removed. Forward and reverse reads were aligned, resulting sequence variant counts (SVs) and taxonomic calls were assigned using the Greengenes 16S rRNA gene database. Models were created, and data were analyzed in R using the following packages: Phyloseq (v. 1.38.0), caret (v. 6.0.92), GGplot2 (v. 3.3.6), mixOmics (6.18.1) and in python using Jupyter notebooks, pandas, scipy, numpy, matplotlib, and seaborn. Venn diagrams were created using the R packages VennDiagram (1.7.3), and ggvenn (0.1.9). Radar plots were created using the R package fsmb (0.7.3). Wilcoxon rank sum tests were used to compare the alpha diversity and genus relative abundances. To calculate statistics, either the Python package Scipy.Stats package or R was used.

### Model generation

The R package randomForest (4.7-1.1) was used to generate a Random Forest (RF) model to classify the samples according to the response to the nutritional intervention and determine important features. Relative SV counts were used to train the RF. The forest used has 1000 trees with a node split (mtry) set to the default of the square root of the number of samples. Biomarkers that were important for classifying patient response to the nutritional intervention at 9 months were also identified using an RF model. Patients with more than one missing test and biomarkers with more than 3 missing patients were removed. After this filtering, we had 148 observations (children) and 23 variables (biomarkers listed in the supplementary file).

O-PLSDA models were created using MATLAB using a custom pipeline initially developed by Remziye Wessel. Lasso regularization is used better to guide the feature selection and model fitting process and improves classification by allowing the selection of a subset of the covariates instead of using all of them. Here, a 5-fold cross-validation was repeated 1000 times to calibrate the model’s performance. Permutation testing results shuffling the labels of the samples show the goodness of fit of the model vs a null distribution, and the cross-validation accuracy showing the stability of the model are reported in supplemental figures. Principal component analysis was performed in R and plotted using ggbiplot (0.55). Correlations between SV abundance and biomarker levels were plotted using corrplot (0.92).

All code and data are available on github (https://github.com/gabehanson/SEEM_microbiome_analysis)

### Validation modeling

Raw sequencing data from Chen et al [13] was processed in R according to the DADA2 pipeline as described above. Change in WAZ was calculated by comparing the WAZ score pre-intervention (day 0 in the original paper) and post-intervention (day 90 in the original paper). Children were ranked according to the change in WAZ and the best 20 and worst 20 responders were compared. Data was transformed into percent composition at the phylum, class, and family levels. These data were imported into Python and combined to form a matrix with percent composition data across the 3 taxonomic levels, and this process was repeated for the SEEM data. Principal component Analysis and PLS modeling were performed using the sklearn package in Python [27].

## Data Availability

All code and data are available on github

https://github.com/gabehanson/SEEM_microbiome_analysis

## Acknowledgments

The authors gratefully acknowledge Carrie Cowardin, Sepideh Dolatshahi, Casey Hoffman, Kevin Janes, Remziye Wessel, Jason Papin, and Greg Medlock for their valuable suggestions and discussion. We also thank the field staff, the data management unit of AKU, and the families and children who participated in the study.

## Author contributions

Conceptualization: Sean Moore, Asad Ali, Junaid Iqbal, Zehra Jamil Methodology: Zehra Jamil, Gabriel Hanson, Brett Moreau Junaid Iqbal

Investigation: Sheraz Ahmed, Fayaz Umrani, Aneeta Hotwani, Furqan Kabir, Kumail Ahmed, Kamran Sadiq, Fatima Aziz

Visualization: Gabriel Hanson, Zehra Jamil Funding acquisition: Asad Ali, Sean Moore

Project administration: Najeeha Iqbal, Sheraz Ahmed, Fayaz Umrani, Aneeta Hotwani, Supervision: Sean Moore, Asad Ali

Writing – original draft: Zehra Jamil, Gabriel Hanson

Writing – review & editing: Sean Moore, Brett Moreau, Indika Mallawaarachchi, Jannie Ma, Najeeha Iqbal, Junaid Iqbal, Asad Ali.

## Table of acronyms

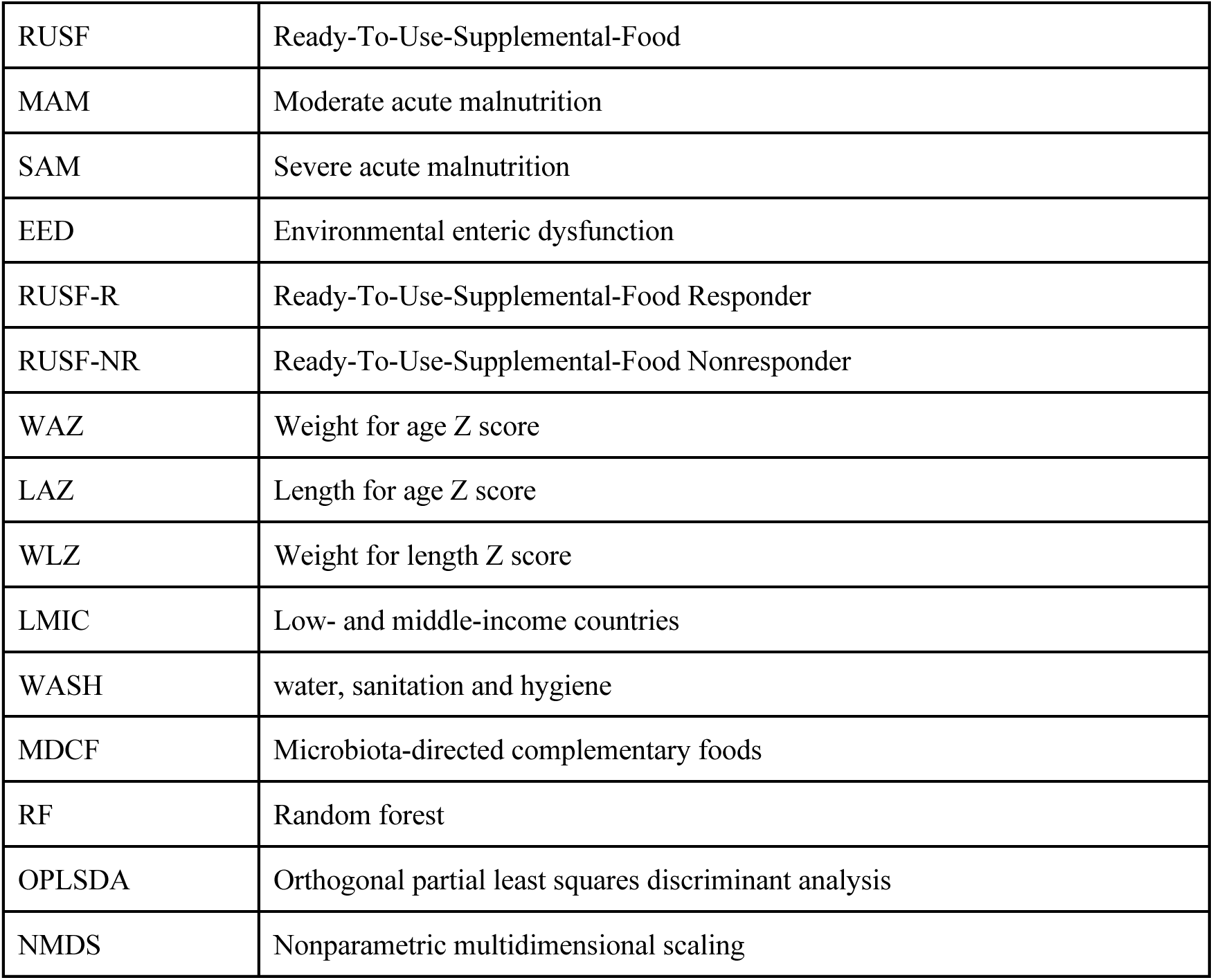
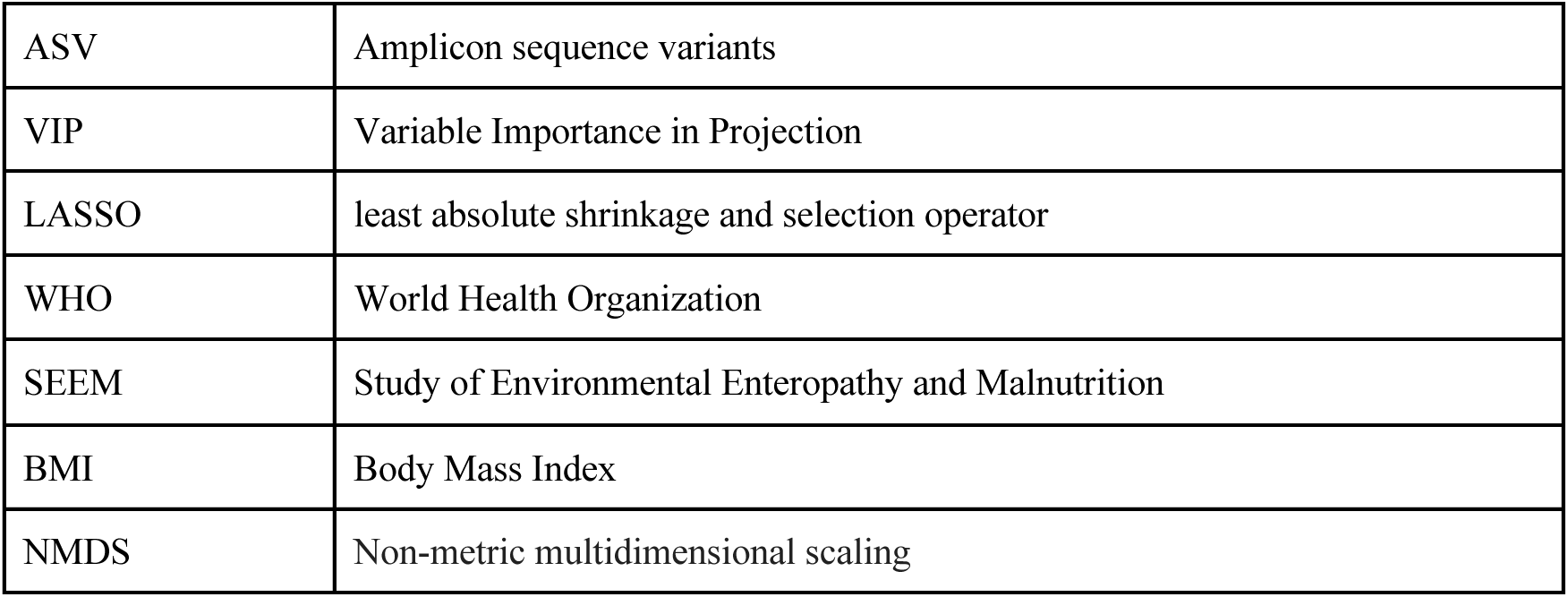

**Table of acronyms:**
|  |  |
| --- | --- |
| RUSF | Ready-To-Use-Supplemental-Food |
| MAM | Moderate acute malnutrition |
| SAM | Severe acute malnutrition |
| EED | Environmental enteric dysfunction |
| RUSF-R | Ready-To-Use-Supplemental-Food Responder |
| RUSF-NR | Ready-To-Use-Supplemental-Food Nonresponder |
| WAZ | Weight for age Z score |
| LAZ | Length for age Z score |
| WLZ | Weight for length Z score |
| LMIC | Low- and middle-income countries |
| WASH | water, sanitation and hygiene |
| MDCF | Microbiota-directed complementary foods |
| RF | Random forest |
| OPLSDA | Orthogonal partial least squares discriminant analysis |
| NMDS | Nonparametric multidimensional scaling |
| ASV | Amplicon sequence variants |
| VIP | Variable Importance in Projection |
| LASSO | least absolute shrinkage and selection operator |
| WHO | World Health Organization |
| SEEM | Study of Environmental Enteropathy and Malnutrition |
| BMI | Body Mass Index |
| NMDS | Non-metric multidimensional scaling |

## List of Supplementary Materials

**Fig S1: Responders and Nonresponders show similar compliance patterns during the nutritional intervention: (A) Adherence to nutritional intervention by the group as shown by the percentage of Acha-mum packets consumed out of the** total packets distributed for each child during the intervention. **(B)** The mean number of vomiting episodes per week of the intervention in each group and error bars show standard deviation. **(C)** Mean number of diarrheal episodes per week of the intervention in each group, error bars show standard deviation.

**Fig S2: PCA of cytokines in SEEM cohort measured at 9 months colored by nutritional status. Arrows show loadings for each variable.** Principal component analysis of serum, urine and fecal biomarker profiles from **(A)** the larger SEEM cohort, wasted children (n=148) and healthy aged-matched children (n=39) and **(B)** wasted cases (n=60) and controls (n=28).

**Fig S3: Additional OPLS-DA figures for Fig 5 (A)** Results of permutation testing for OPLSDA model in figure 5. The plot shows a histogram of cross-validation accuracy for 1000 randomly permuted models. The correctly labeled model is shown as a red star. **(B)** Jitter plots showing ASV counts per sample for VIPs identified in OPLSDA analysis. **(C)** The heat map shows a correlation between important ASVs identified in OPLS-DA and other ASVs with >70% correlation, which were removed as linearly correlated. The X-axis has only important features labeled, while the y-axis has correlated, and important features labeled.

**Fig S4: Additional OPLS-DA figures for Fig. 6 (A)** Results of permutation testing for OPLSDA model in figure 5. Plot shows a histogram of cross validation accuracy for 1000 randomly permuted models. Correctly labeled model is shown as a red star. **(B)** Jitter plots showing ASV counts per sample for VIPs identified in OPLSDA analysis. **(C)** Heatmap showing a correlation between important ASVs identified in OPLS-DA and other ASVs with >70% correlation that was removed as linearly correlated. The X-axis has only important features labeled, while the y-axis has correlated and important features.

**Fig S5: Microbiome shifts during intervention for Fig. 7 (A)** Alpha diversity of control samples at 9 months and 12 months of life. **(B)** Bar plot showing average relative abundance at Phylum level in responders and nonresponders pre and post-intervention. **(C)** Bar plot showing average relative abundance at a class level in responders and nonresponders pre- and post-intervention.

**Fig S6: Additional OPLS-DA figures for Fig. 7 (A)** Results of permutation testing for OPLSDA model in figure 5. The plot shows a histogram of cross-validation accuracy for 1000 randomly permuted models. The correctly labeled model is shown as a red star. **(B)** Jitter plots showing ASV counts per sample for VIPs identified in OPLSDA analysis. **(C)** Correlation network of ASVs with >70% correlation of ASV identified in OPLS-DA. The X-axis has only important features labeled, while the y-axis has correlated and important features labeled.

**Fig S7: Inflamed nonresponders show similar patterns of adherence and diarrheal episodes but increased respiratory infections: (A)** Adherence to nutritional intervention by group as shown by the percentage of Acha Mum packets consumed out of total packets distributed for each child during the intervention. **(B)** Mean number of diarrheal episodes per week of the intervention in each group, error bars show standard deviation.

**Fig S8: Separate cohort of malnourished children shows different trends in microbiome restructuring during the intervention: (A)** Children from the validation dataset who received the chickpea-based intervention were sorted by change in WAZ during the intervention, and the 20 best and 20 worst responders were compared. **(B)** Bar plots of microbiome composition at pre and post-intervention timepoints in responder and nonresponder groups. **(C)** Scatter plot of the X scores on latent variables 1 and 2 (LV1 & LV2), where each point represents one sample. **(D)** Principal component analysis of combined family, class and phylum level microbiome composition data. **(E)** The PLS model was trained on SEEM data and tested on Chen et al. data. Confusion matrix and ROC curves describe the accuracy of the model.

---

**Supplementary Table 1:**
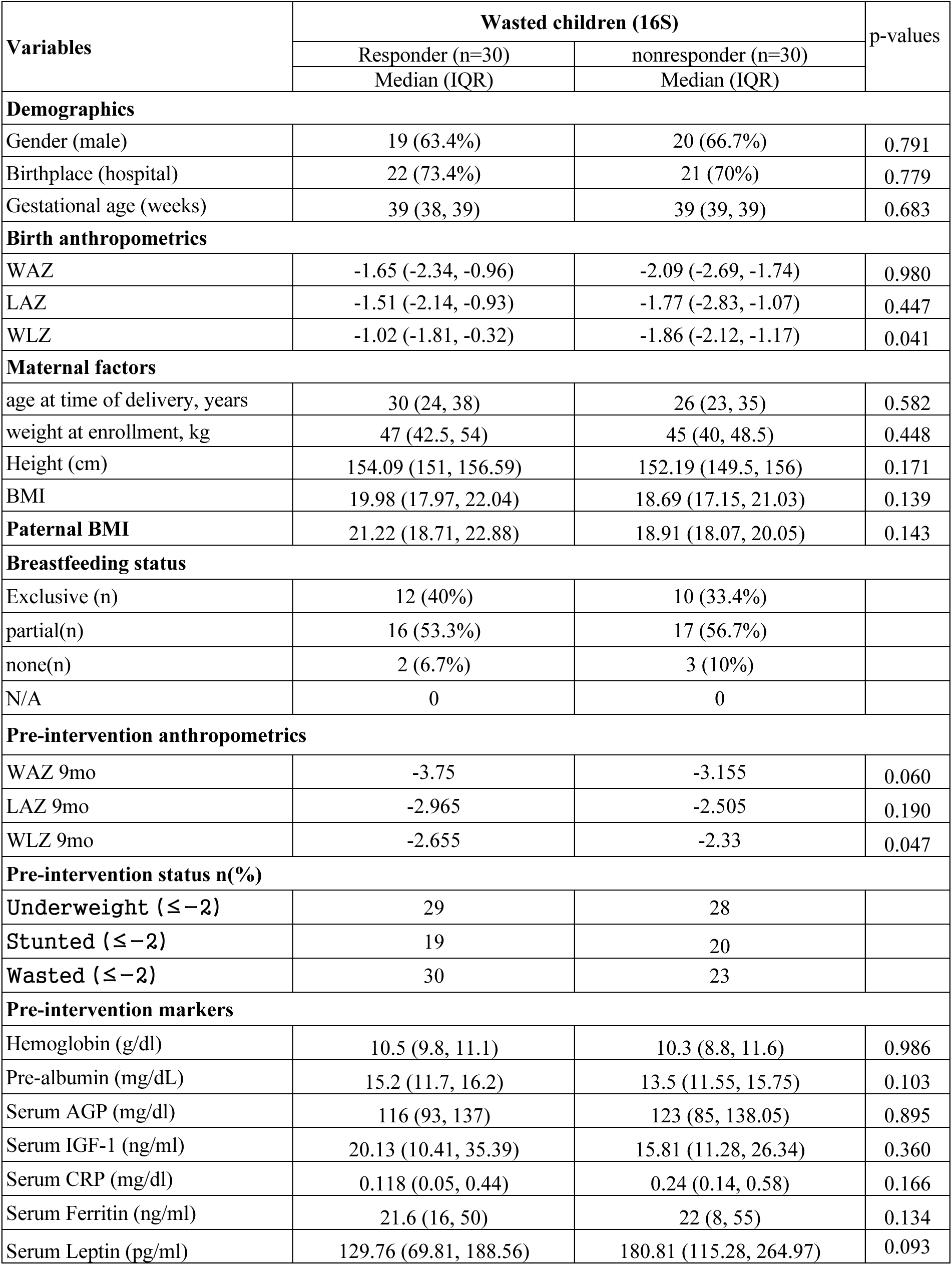
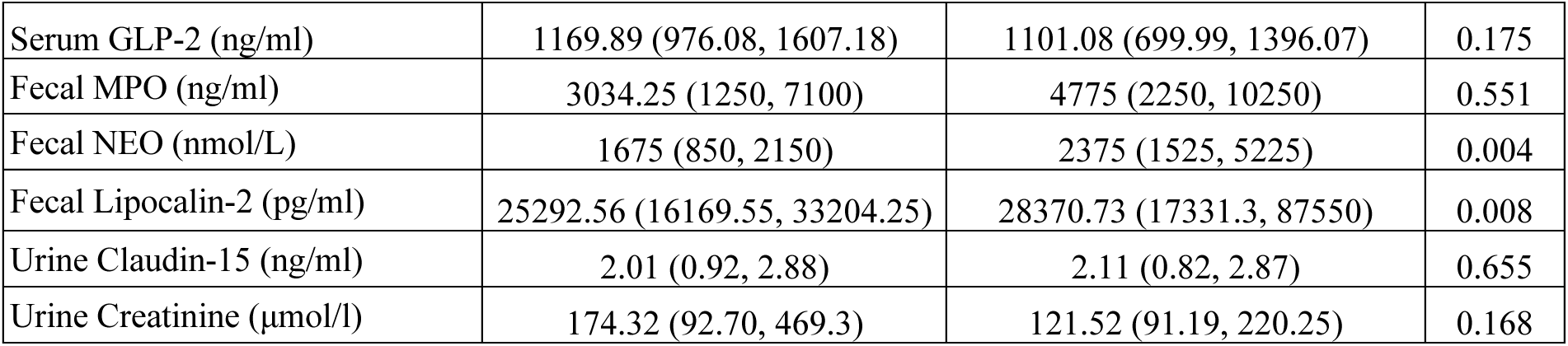
Baseline characteristics of cases only (30 best and 30 worst responders).

| Variables | Wasted children (16S) |  | p-values |
| --- | --- | --- | --- |
|  | Responder (n=30) | nonresponder (n=30) |  |
|  | Median (IQR) | Median (IQR) |  |
| Demographics |  |  |  |
| Gender (male) | 19 (63.4%) | 20 (66.7%) | 0.791 |
| Birthplace (hospital) | 22 (73.4%) | 21 (70%) | 0.779 |
| Gestational age (weeks) | 39 (38, 39) | 39 (39, 39) | 0.683 |
| Birth anthropometrics |  |  |  |
| WAZ | -1.65 (-2.34, -0.96) | -2.09 (-2.69, -1.74) | 0.980 |
| LAZ | -1.51 (-2.14, -0.93) | -1.77 (-2.83, -1.07) | 0.447 |
| WLZ | -1.02 (-1.81, -0.32) | -1.86 (-2.12, -1.17) | 0.041 |
| Maternal factors |  |  |  |
| age at time of delivery, years | 30 (24, 38) | 26 (23, 35) | 0.582 |
| weight at enrollment, kg | 47 (42.5, 54) | 45 (40, 48.5) | 0.448 |
| Height (cm) | 154.09 (151, 156.59) | 152.19 (149.5, 156) | 0.171 |
| BMI | 19.98 (17.97, 22.04) | 18.69 (17.15, 21.03) | 0.139 |
| Paternal BMI | 21.22 (18.71, 22.88) | 18.91 (18.07, 20.05) | 0.143 |
| Breastfeeding status |  |  |  |
| Exclusive (n) | 12 (40%) | 10 (33.4%) |  |
| partial(n) | 16 (53.3%) | 17 (56.7%) |  |
| none(n) | 2 (6.7%) | 3 (10%) |  |
| N/A | 0 | 0 |  |
| Pre-intervention anthropometrics |  |  |  |
| WAZ 9mo | -3.75 | -3.155 | 0.060 |
| LAZ 9mo | -2.965 | -2.505 | 0.190 |
| WLZ 9mo | -2.655 | -2.33 | 0.047 |
| Pre-intervention status n(%) |  |  |  |
| Underweight ($\leq -2$) | 29 | 28 | |
| Stunted ($\leq -2$) | 19 | 20 | |
| Wasted ($\leq -2$) | 30 | 23 | |
| Pre-intervention markers |  |  |  |
| Hemoglobin (g/dl) | 10.5 (9.8, 11.1) | 10.3 (8.8, 11.6) | 0.986 |
| Pre-albumin (mg/dL) | 15.2 (11.7, 16.2) | 13.5 (11.55, 15.75) | 0.103 |
| Serum AGP (mg/dl) | 116 (93, 137) | 123 (85, 138.05) | 0.895 |
| Serum IGF-1 (ng/ml) | 20.13 (10.41, 35.39) | 15.81 (11.28, 26.34) | 0.360 |
| Serum CRP (mg/dl) | 0.118 (0.05, 0.44) | 0.24 (0.14, 0.58) | 0.166 |
| Serum Ferritin (ng/ml) | 21.6 (16, 50) | 22 (8, 55) | 0.134 |
| Serum Leptin (pg/ml) | 129.76 (69.81, 188.56) | 180.81 (115.28, 264.97) | 0.093 |
| Serum GLP-2 (ng/ml) | 1169.89 (976.08, 1607.18) | 1101.08 (699.99, 1396.07) | 0.175 |
| Fecal MPO (ng/ml) | 3034.25 (1250, 7100) | 4775 (2250, 10250) | 0.551 |
| Fecal NEO (nmol/L) | 1675 (850, 2150) | 2375 (1525, 5225) | 0.004 |
| Fecal Lipocalin-2 (pg/ml) | 25292.56 (16169.55, 33204.25) | 28370.73 (17331.3, 87550) | 0.008 |
| Urine Claudin-15 (ng/ml) | 2.01 (0.92, 2.88) | 2.11 (0.82, 2.87) | 0.655 |
| Urine Creatinine (μmol/l) | 174.32 (92.70, 469.3) | 121.52 (91.19, 220.25) | 0.168 |

