## Supplemental Material for "Host-microbiome determinants of ready-to-use supplemental food efficacy in acute childhood malnutrition"

### Supplementary Material and Methods

**Study participant details:** Study participants were drawn from the SEEM-Pakistan birth cohort study (N=416) conducted in Matiari, Sindh. We selected participants for microbiome analysis based on the availability of longitudinally collected fecal samples (S Fig 1). SEEM-Pakistan explored gut tissue histology, transcriptomics, and intestinal permeability in undernourished cases refractory to nutritional interventions. The study was approved by the Aga Khan University Ethical Review Committee (ERC # 2021-0535-19973), and written informed consent was obtained from the participant's parents. The study was registered at ClinicalTrials.gov ID NCT03588013, and the SEEM protocols were published [10].

Newborns were enrolled at 0-3 months of age and followed monthly for growth monitoring. We monitored their weight and length monthly for up to six months of age and then classified study participants as either well-nourished controls (WLZ ≥0, LAZ ≥-1, n = 51) or undernourished cases (wasted with WLZ ≤-2, n = 365) based on two consecutive months of anthropometry (**Fig 1**).

### Supplementary Results

Baseline clinical characteristics for case (n=60) and control (n=28) infants for whom complete biomarker and fecal 16S microbiome data were available are summarized in **S Table 1**. Maternal and paternal BMI as well as birth anthropometrics were significantly lower in cases vs. controls (P <0.05), differences that became even more pronounced by nine months of age (P <0.05). Breastfeeding patterns were similar between groups over the first six months of life. Cases in which complete biomarker data were available broadly represented the larger SEEM cohort, as shown in **S Table 2**. The comparison between the baseline characteristics of the larger SEEM cohort and those selected for 16S fecal microbiome analysis is summarized in S Table 3, reporting these selected cases and controls as true representatives of their respective larger groups.

S Table 1: Comparison of baseline characteristics between cases (responders and nonresponders) and controls (n=88)

| Variables | Wasted cases n=60 | Controls n=28 | P-values |
|---|---|---|---|
| Demographics | | | |
| Gender (males) | 39 (65%) | 18 (64.3%) | 0.940 |
| Birthplace (hospital) | 43 (71.6%) | 24 (85.7%) | 0.120 |
| Gestational age- median (IQR) | 39 (38, 39) | 39 (39, 39) | 0.054 |
| Anthropometrics at birth (median IQR) | | | |
| Weight-for-age z score | -1.99 (-2.69, -1.32) | -1.03 (-1.39, -0.31) | < 0.001 |
| Length-for-age age z score | -1.59 (-2.51, -1.06) | -0.88 (-1.54, -0.16) | < 0.001 |
| Weight-for-length z score | -1.39 (-1.99, -0.72) | -0.45 (-1.16, -0.09) | < 0.001 |
| Maternal factors (median IQR) | | | |
| Age | 27 (23.5, 35) | 27 (25, 30) | 0.263 |
| Body mass index | 19.44 (17.63, 21.89) | 20.92 (19.76, 24.46) | 0.045 |
| Paternal BMI (median IQR) | 19.39 (18.47, 21.89) | 22.46 (20.27, 24.32) | 0.012 |
| Breastfeeding status (%) | | | |
| Exclusive | 22 (36.7%) | 10 (35.7%) | 0.238 |
| Partial | 33 (55%) | 15 (53.6%) | 0.238 |
| None | 5 (8.3%) | 1 (3.3%) | 0.238 |
| Anthropometrics at age 9 months | | | |
| Weight-for-age z score | -3.47 (-4.10, -2.67) | -0.11 ( -0.72, 0.27) | <0.001 |
| Length-for-age age z score | -2.59 (3.55, -1.75) | -0.88 ( -1.88, -0.06) | <0.001 |
| Weight-for-length z score | -2.56 (-3.09, -2.12) | 0.53 (-0.17, 1.12) | <0.001 |

Supplementary Table 2: Baseline characteristics compared between cases only (30 best and 30 worst responders).

| Variables | Wasted children (16S) | | p-values |
|---|---|---|---|
| | Responder (n=30) Median (IQR) | nonresponder (n=30) Median (IQR) | |
| **Demographics** | | | |
| Gender (male) | 19 (63.4%) | 20 (66.7%) | 0.791 |
| Birthplace (hospital) | 22 (73.4%) | 21 (70%) | 0.779 |
| Gestational age (weeks) | 39 (38, 39) | 39 (39, 39) | 0.683 |
| **Birth anthropometrics** | | | |
| WAZ | -1.65 (-2.34, -0.96) | -2.09 (-2.69, -1.74) | 0.980 |
| LAZ | -1.51 (-2.14, -0.93) | -1.77 (-2.83, -1.07) | 0.447 |
| WLZ | -1.02 (-1.81, -0.32) | -1.86 (-2.12, -1.17) | 0.041 |
| **Maternal factors** | | | |
| age at time of delivery, years | 30 (24, 38) | 26 (23, 35) | 0.582 |
| weight at enrollment, kg | 47 (42.5, 54) | 45 (40, 48.5) | 0.448 |
| Height (cm) | 154.09 (151, 156.59) | 152.19 (149.5, 156) | 0.171 |
| BMI | 19.98 (17.97, 22.04) | 18.69 (17.15, 21.03) | 0.139 |
| **Paternal BMI** | 21.22 (18.71, 22.88) | 18.91 (18.07, 20.05) | 0.143 |
| **Breastfeeding status** | | | |
| Exclusive (n) | 12 (40%) | 10 (33.4%) | |
| partial(n) | 16 (53.3%) | 17 (56.7%) | |
| none(n) | 2 (6.7%) | 3 (10%) | |
| N/A | 0 | 0 | |
| **Pre-intervention anthropometrics** | | | |
| WAZ 9mo | -3.75 | -3.155 | 0.060 |
| LAZ 9mo | -2.965 | -2.505 | 0.190 |
| WLZ 9mo | -2.655 | -2.33 | 0.047 |
| **Pre-intervention status n(%)** | | | |
| Underweight (≤-2) | 29 | 28 | |
| Stunted (≤-2) | 19 | 20 | |
| Wasted (≤-2) | 30 | 23 | |
| **Pre-intervention markers** | | | |
| Hemoglobin (g/dl) | 10.5 (9.8, 11.1) | 10.3 (8.8, 11.6) | 0.986 |
| Pre-albumin (mg/dL) | 15.2 (11.7, 16.2) | 13.5 (11.55, 15.75) | 0.103 |
| Serum AGP (mg/dl) | 116 (93, 137) | 123 (85, 138.05) | 0.895 |
| Serum IGF-1 (ng/ml) | 20.13 (10.41, 35.39) | 15.81 (11.28, 26.34) | 0.360 |
| Serum CRP (mg/dl) | 0.118 (0.05, 0.44) | 0.24 (0.14, 0.58) | 0.166 |
| Serum Ferritin (ng/ml) | 21.6 (16, 50) | 22 (8, 55) | 0.134 |
| Serum Leptin (pg/ml) | 129.76 (69.81, 188.56) | 180.81 (115.28, 264.97) | 0.093 |
| Serum GLP-2 (ng/ml) | 1169.89 (976.08, 1607.18) | 1101.08 (699.99, 1396.07) | 0.175 |

| | | | |
|---|---|---|---|
| Fecal MPO (ng/ml) | 3034.25 (1250, 7100) | 4775 (2250, 10250) | 0.551 |
| Fecal NEO (nmol/L) | 1675 (850, 2150) | 2375 (1525, 5225) | 0.004 |
| Fecal Lipocalin-2 (pg/ml) | 25292.56 (16169.55, 33204.25) | 28370.73 (17331.3, 87550) | 0.008 |
| Urine Claudin-15 (ng/ml) | 2.01 (0.92, 2.88) | 2.11 (0.82, 2.87) | 0.655 |
| Urine Creatinine (μmol/l) | 174.32 (92.70, 469.3) | 121.52 (91.19, 220.25) | 0.168 |

**Supplementary Table 3:** Comparison between SEEM cohort and selected cases for this sub-study: either here on via excel sheet

| Variables | All Wasted children (n=186) receiving Nutritional Intervention | | | | | | Controls | | |
|---|---|---|---|---|---|---|---|---|---|
| | Responders | | P | Non-responder | | p | | | p |
| | responder in SEEM | with fecal 16S data | | non-responder in SEEM | with fecal 16S data | | controls in SEEM | with fecal 16S data | |
| **N** | 87 | 30 | | 99 | 30 | | 48 | 28 | |
| **Demographics** | | | | | | | | | |
| Gender | 59 | 19 | 0.66 | 58 | 20 | 0.42 | 27 | 18 | 0.49 |
| Gest age | 39 (39 39) | 39 (38 39) | 0.63 | 39 (39 39) | 39 (39 39) | 0.16 | 39 (39 39) | 39 (39 39) | 0.44 |
| **Birth anthropometrics** | | | | | | | | | |
| WAZ | -2.2 (-3.17,-1.33) | -1.645 (-2.34, -0.96) | 0.06 | -2.21 (-2.795, -1.655) | -2.09 (-2.69, -1.74) | 0.98 | -1.03 (-1.585, -0.39) | -1.03 (-1.385, -0.305) | 0.75 |
| LAZ | -2.13 (-2.94, -1.15) | -1.51 (-2.14, -0.93) | 0.03 | -1.83 (-2.63, -1.195) | -1.765 (-2.83, -1.07) | 0.96 | -0.895 (-1.56, -0.065) | -0.88 (-1.54, -0.16) | 0.66 |
| WLZ | -1.26 (-1.96, -0.48) | -1.02 (-1.81, -0.32) | 0.52 | -1.59 (-2.15, -0.89) | -1.855 (-2.12, -1.17) | 0.59 | -0.48 (-1.13, -0.095) | -0.445 (-1.155, -0.095) | 1.00 |
| **Maternal factors** | | | | | | | | | |
| age | 29 (24, 35) | 30 (24, 38) | 0.44 | 26 (23, 34) | 26(23, 35) | 0.65 | 26 (23, 30) | 27 (25, 30) | 0.65 |
| weight | 46 (41.7,51.5) | 47 (42.5, 54) | 0.42 | 44.5 (40, 48.79) | 45 (40, 48.5) | 0.62 | 49.54 (44.3,53.8) | 49.25 (44.5, 53.60) | 0.69 |
| height | 152.39 (149.75, 155.34) | 154.09 (151, 156.58) | 0.05 | 151.80 (147.94, 155) | 152.19 (149.5, 156) | 0.14 | 152.05 (148.34, 157) | 151.94 (148.25, 155.54) | 0.47 |
| BMI | 19.73 (18.5,21.7) | 19.97 (17.9,22.42) | 0.73 | 19.35 (17.52, 21.3) | 18.68 (17.154, 21.02) | 0.43 | 20.99 (19.5,24.28) | 20.92 (19.76, 24.46) | 0.88 |
| **Paternal BMI** | 19.90 (18.71, 22.37) | 21.21 (18.71, 22.87) | 0.90 | 19.34 (18.03, 21.75) | 18.91 (18.06, 20.04) | 0.45 | 21.85 (19.85, 25.17) | 22.45 (20.27, 24.317) | 0.73 |
| **Breast feeding status** | | | | | | | | | |
| exclusive | 38 | 12 | | 34 | 10 | | 16 | 10 | |
| partial | 45 | 16 | | 54 | 17 | | 28 | 15 | |
| none | 4 | 2 | | 11 | 3 | | 3 | 2 | |
| **Pre-intervention anthropometrics** | | | | | | | | | |

| | | | | | | | | | |
|---|---|---|---|---|---|---|---|---|---|
| WAZ 9mo | -3.65 | -3.75 | 0.89 | -3.28 | -3.155 | 0.76 | -0.005 | -0.11 | 0.41 |
| LAZ 9mo | -2.89 | -2.965 | 0.96 | -2.49 | -2.505 | 0.67 | -0.9 | -0.88 | 0.79 |
| WLZ 9mo | -2.62 | -2.655 | 0.72 | -2.37 | -2.33 | 0.76 | 0.595 | 0.53 | 0.59 |
| **Pre-intervention status n(%)** | | | | | | | | | |
| Wasted | 84 | 29 | | 94 | 28 | | 2 | 0 | |
| Underweight | 61 | 19 | | 63 | 20 | | 8 | 5 | |
| Stunted | 81 | 30 | | 82 | 23 | | 0 | 0 | |
| **Pre-intervention markers** | | | | | | | | | |
| Hb | 10.4 (9.3 ,11.4) | 10.5 (9.8, 11.1) | 0.95 | 10.4 (8.9, 11.2) | 10.3 (8.8, 11.6) | 0.91 | 10.7 (10.15, 11.7) | 10.45 (9.5, 11.3) | 0.46 |
| Pre-albumin | 15.2 (12.9, 16.6) | 15.2 (11.7, 16.2) | 0.50 | 13.2 (11.55, 15.75) | 13.5 (11.55, 15.75) | 0.66 | 15.3 (13.7, 17.7) | 15.2 (13.6, 16.35) | 0.17 |
| Serum AGP mg/dl | 110.5 (82 ,142.4) | 116 (93, 137) | 0.58 | 106.5 (78.5, 138.44) | 123 (85, 138.05) | 0.65 | 94.765 (72, 126) | 115 (85, 134) | 0.16 |
| Serum IGF1 ng/ml | 19.06 (10.48, 29.44) | 20.13 (10.41, 35.39) | 0.56 | 15.81 (8.87, 24.52) | 15.81 (11.28, 26.34) | 0.94 | 27.345 (19.26, 37.65) | 24.46 (17.57, 36.48) | 0.52 |
| Serum CRP | 0.136 (0.048, 0.39) | 0.118 (0.045, 0.44) | 0.62 | 0.24 (0.115, 0.64) | 0.24 (0.14, 0.58) | 0.94 | 0.0945 (0.05, 0.23) | 0.1295 (0.066, 0.368) | 0.88 |
| Serum Ferritin | 20.5 (7.2, 42.5) | 21.6 (16, 50) | 0.38 | 19 8 (05, 39.5) | 22 (8, 55) | 0.83 | 9.95 (5.5, 22) | 16.95 (5.8, 34.8) | 0.36 |
| Fecal MPO ng/ml | 2858.25 (1050, 6000) | 3034.25 (1250, 7100) | 0.59 | 4933 (1850, 11600) | 4775 (2250, 10250) | 0.19 | 4672.25 (2079.5, 10575) | 4672.25 (2640.25, 9275) | 0.75 |
| Fecal NEO nmol/L | 1482.75 (627.5, 2350) | 1675 (850, 2150) | 0.96 | 2037.5 (1228.5, 3651.25) | 2375 (1525, 5225) | 0.20 | 1633.5 (630, 2750) | 1341.5 (485.95, 2600) | 0.96 |
| Fecal Lipocalin | 20526.15 (12647.85, 32778.5) | 25292.58 (16169.55, 33204.25) | 0.74 | 24134.53 (15475.5, 35537) | 28370.73 (17331.3, 87550) | 0.04 | 27370 (17347.08, 67500) | 40097.925 (20119.5, 94775) | 0.29 |
| Urine claudin-15 | 1.783 (0.93,2.87) | 2.01 (0.91,2.88) | 0.57 | 1.4 (0.72,2.66) | 2.1065 (0.81,2.86) | 0.46 | 0.909 (0.72,1.13) | 0.924 (0.68, 1.15) | 0.78 |
| Urine creatinine | 139.36 (88.38, 287.24) | 174.32 (92.70, 469.3) | 0.32 | 113.80 (68.95, 212.04) | 121.51 (91.19, 220.25) | 0.61 | 163.79 (107.98, 205.83) | 176.57 (129.65, 212.41) | 0.54 |
| Serum Leptin | 122.5 (73.22,217.48) | 129.76 (69.81, 188.56) | 0.57 | 140.03 (83.68, 217.91) | 180.81 (115.28, 264.97) | 0.21 | 279.85 (190.79, 389.19) | 258.17 (179.39, 345.51) | 0.38 |
| Serum GLP | 1288.34 (908.43, 1747.313) | 1169.89 (976.08, 1607.18) | 0.58 | 993.43 (723.63, 1350.44) | 1101.08 (699.99, 1396.07) | 0.95 | 1705.90 (843.92, 2688.24) | 1260.87 (649.51, 2286.14) | 0.42 |

Fig S1b

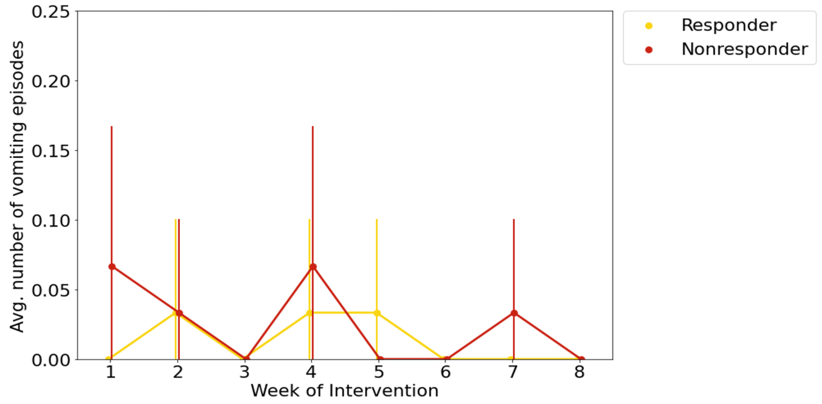


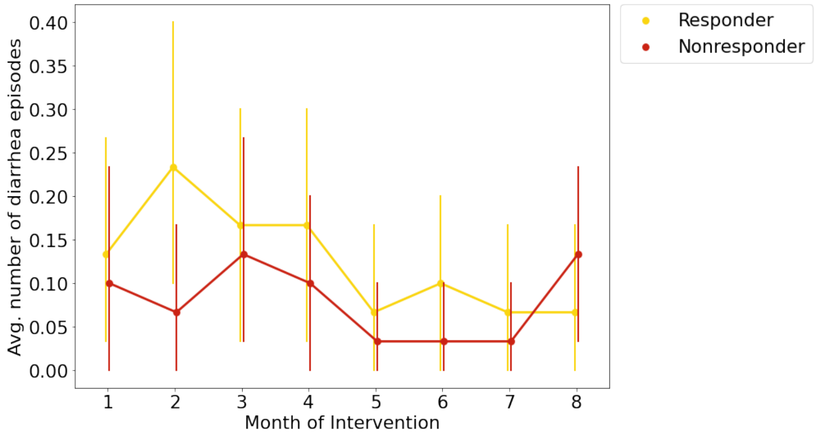


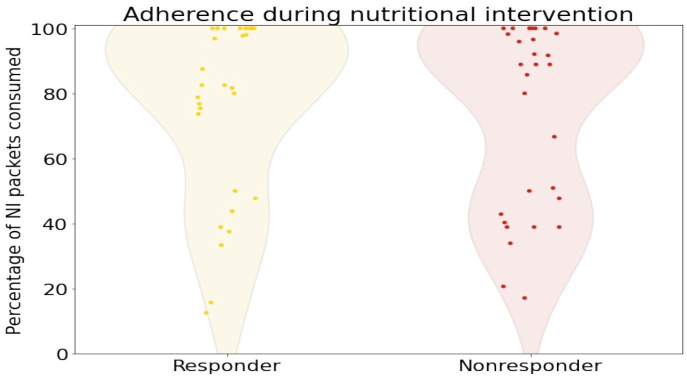


**Fig S1: Responders and Nonresponders show similar compliance patterns during the nutritional intervention: (A) Adherence to nutritional intervention by the group as shown by the percentage of Acha-mum packets consumed out of the** total packets distributed for each child during the intervention. **(B)** The mean number of vomiting episodes per week of the intervention in each group and error bars show standard deviation. **(C)** Mean number of diarrheal episodes per week of the intervention in each group, error bars show standard deviation.

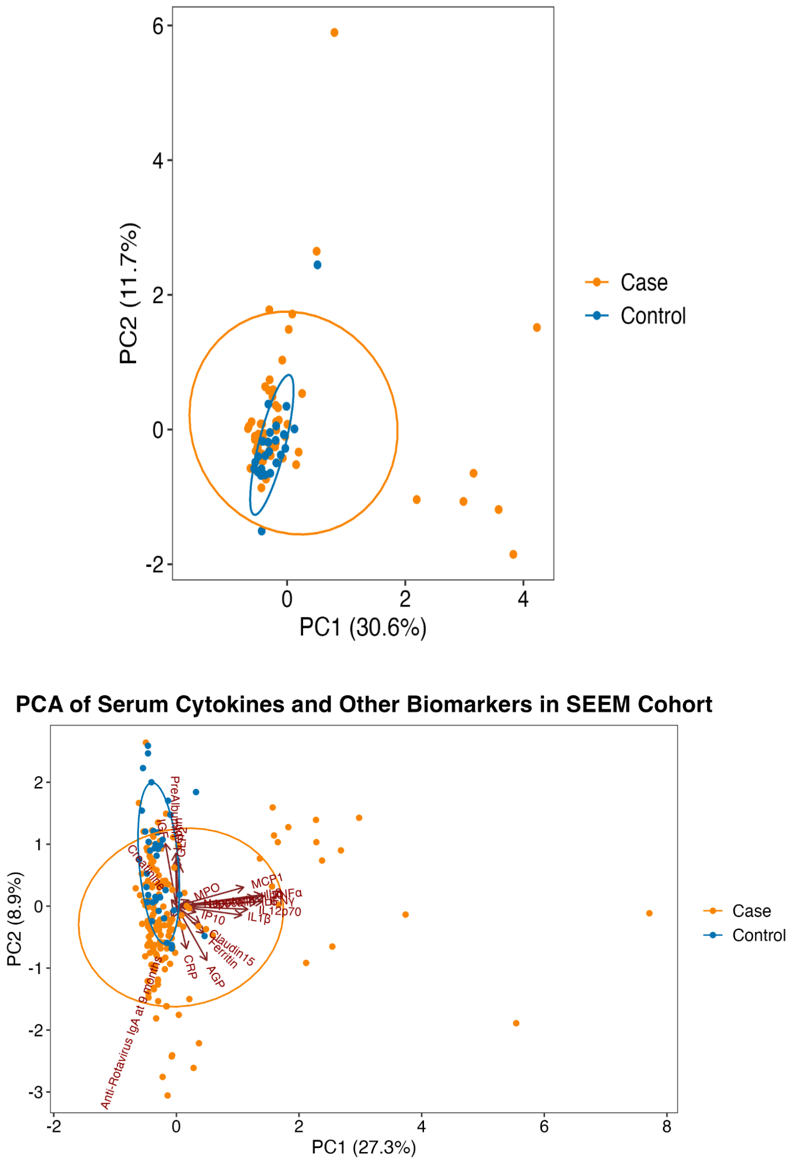


**Fig S2: PCA of cytokines in SEEM cohort measured at 9 months colored by nutritional status. Arrows show loadings for each variable.** Principal component analysis of serum, urine and fecal biomarker profiles from **(A)** the larger SEEM cohort, wasted children (n=148) and healthy aged-matched children (n=39) and **(B)** wasted cases (n=60) and controls (n=28).

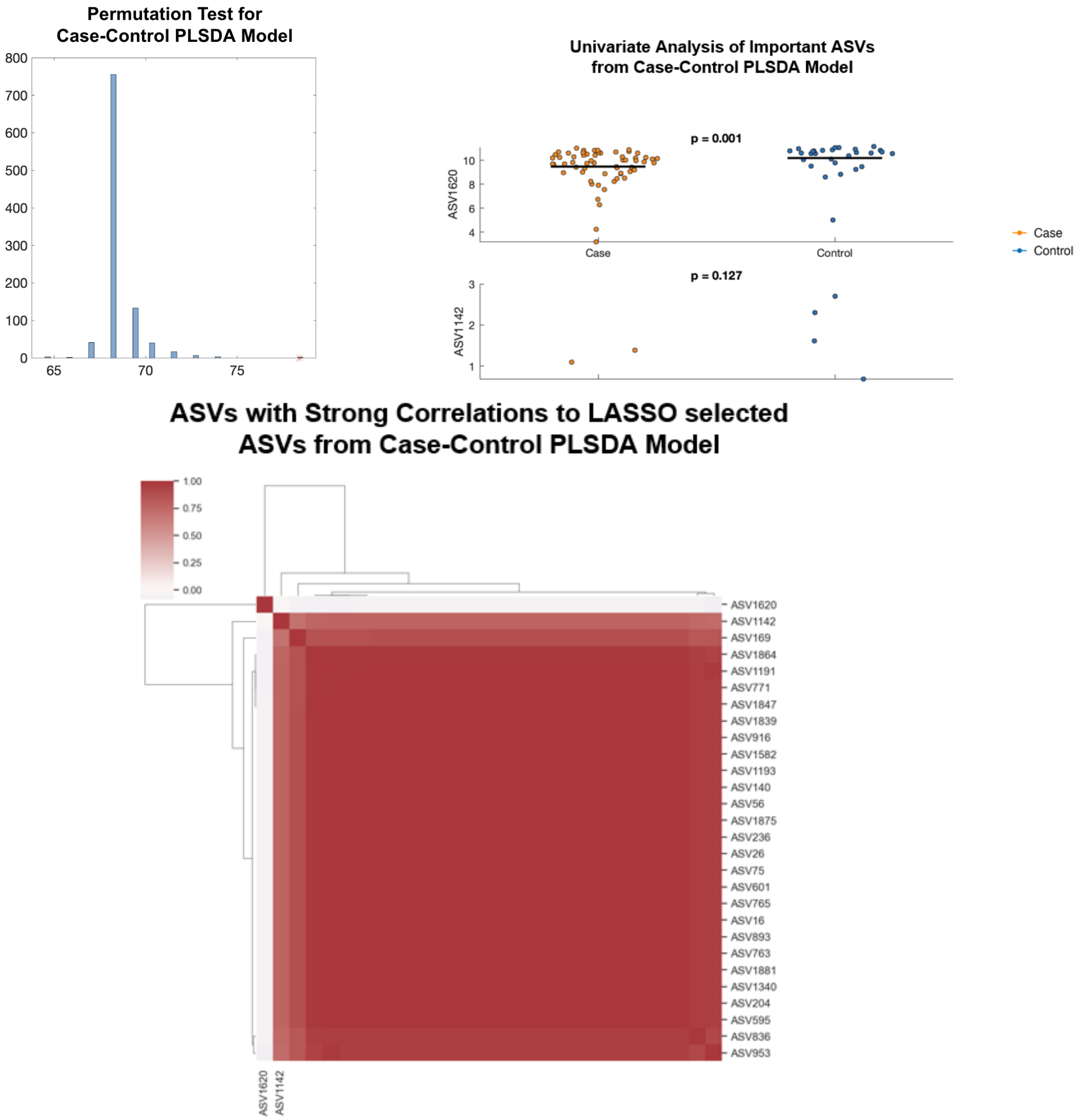


**Fig S3: Additional OPLS-DA figures for Fig 5 (A)** Results of permutation testing for OPLSDA model in figure 5. The plot shows a histogram of cross-validation accuracy for 1000 randomly permuted models. The correctly labeled model is shown as a red star. **(B)** Jitter plots showing ASV counts per sample for VIPs identified in OPLSDA analysis. **(C)** The heat map shows a correlation between important ASVs identified in OPLS-DA and other ASVs with >70% correlation, which were removed as linearly correlated. The X-axis has only important features labeled, while the y-axis has correlated, and important features labeled.

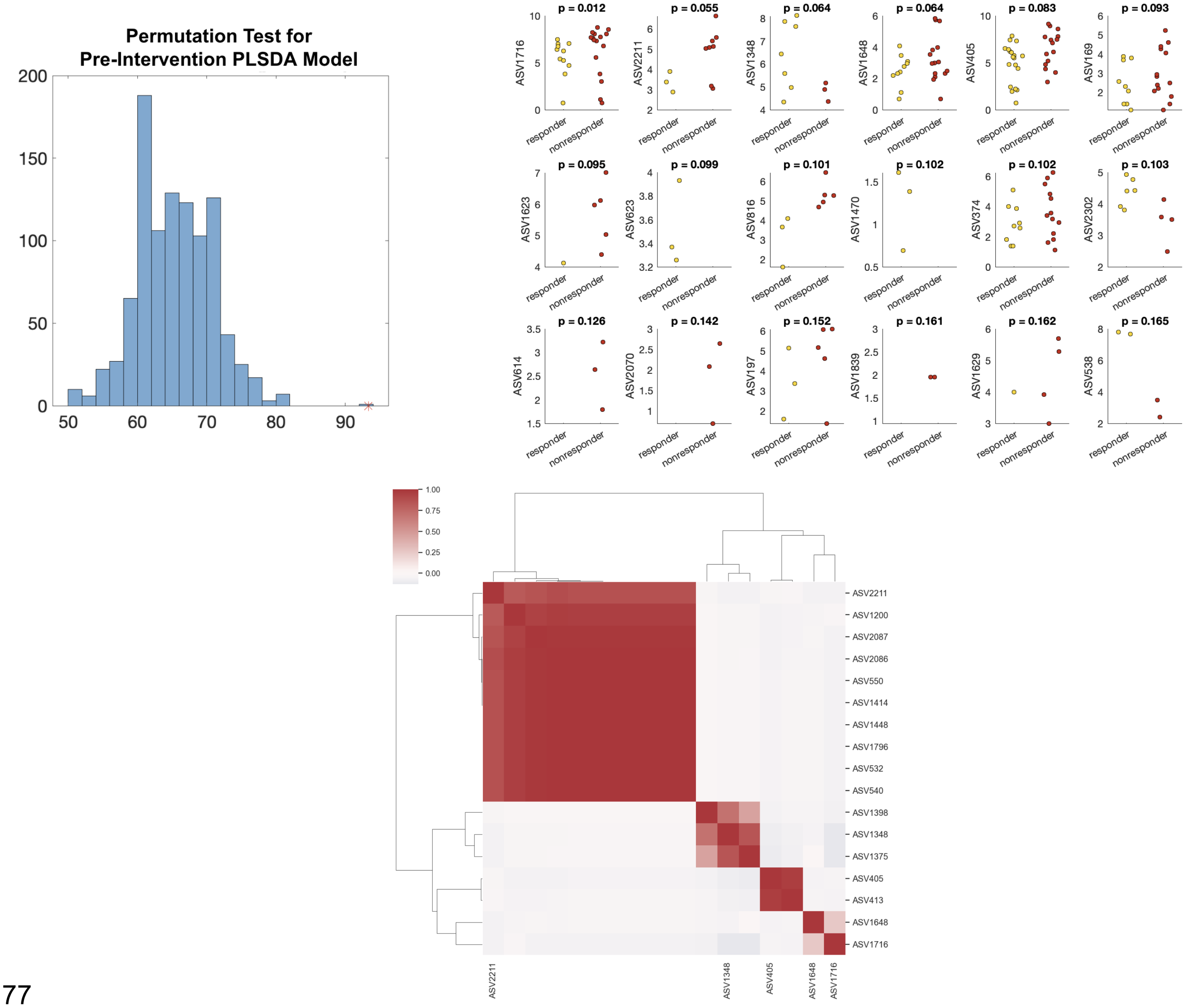


**Fig S4: Additional OPLS-DA figures for Fig. 6 (A)** Results of permutation testing for OPLSDA model in figure 5. Plot shows a histogram of cross validation accuracy for 1000 randomly permuted models. Correctly labeled model is shown as a red star. **(B)** Jitter plots showing ASV counts per sample for VIPs identified in OPLSDA analysis. **(C)** Heatmap showing a correlation between important ASVs identified in OPLS-DA and other ASVs with >70% correlation that was removed as linearly correlated. The X-axis has only important features labeled, while the y-axis has correlated and important features.

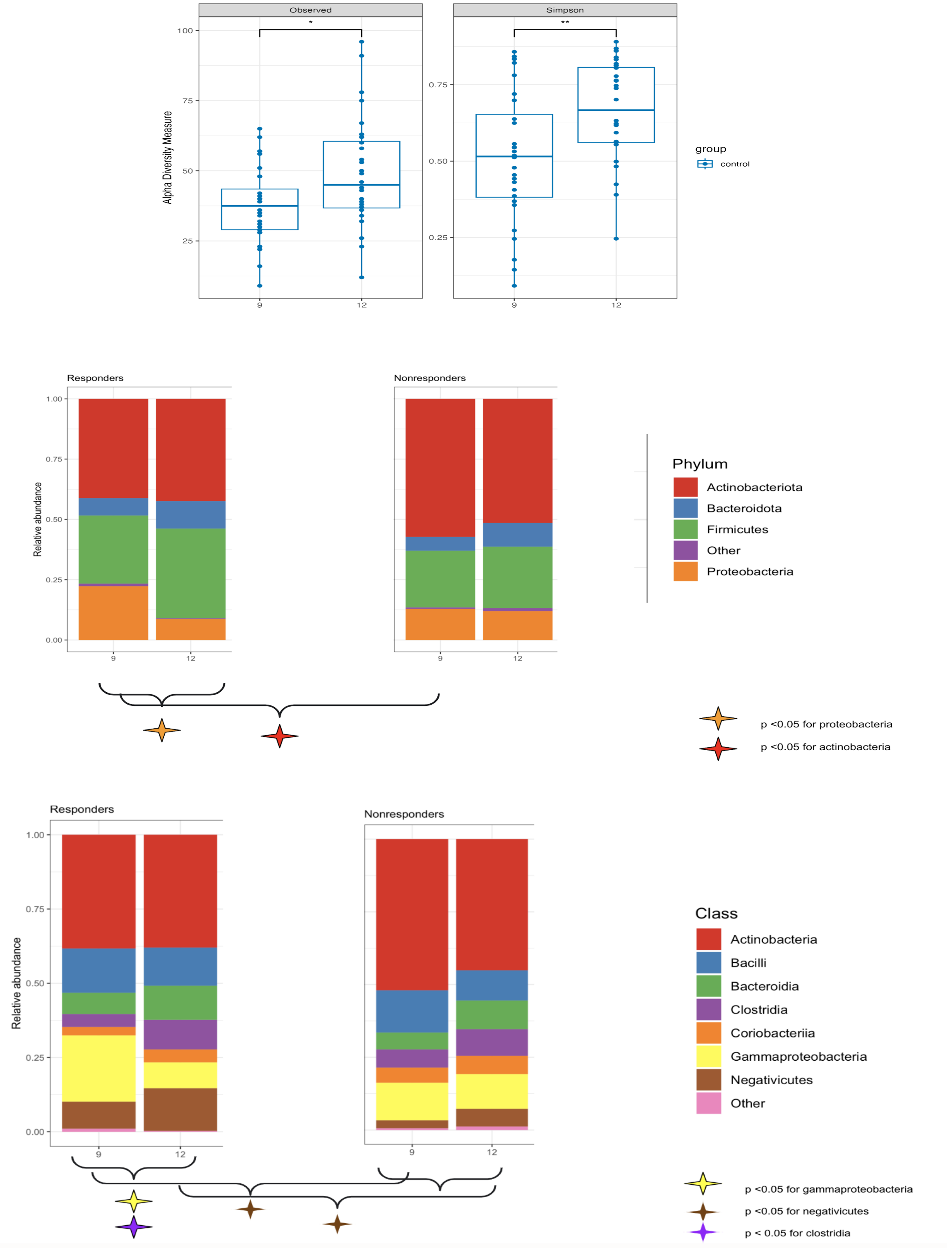


**Fig S5: Microbiome shifts during intervention for Fig. 7 (A)** Alpha diversity of control samples at 9 months and 12 months of life. **(B)** Bar plot showing average relative abundance at Phylum level in responders and nonresponders pre and post-intervention. **(C)** Bar plot showing average relative abundance at a class level in responders and nonresponders pre- and post-intervention.

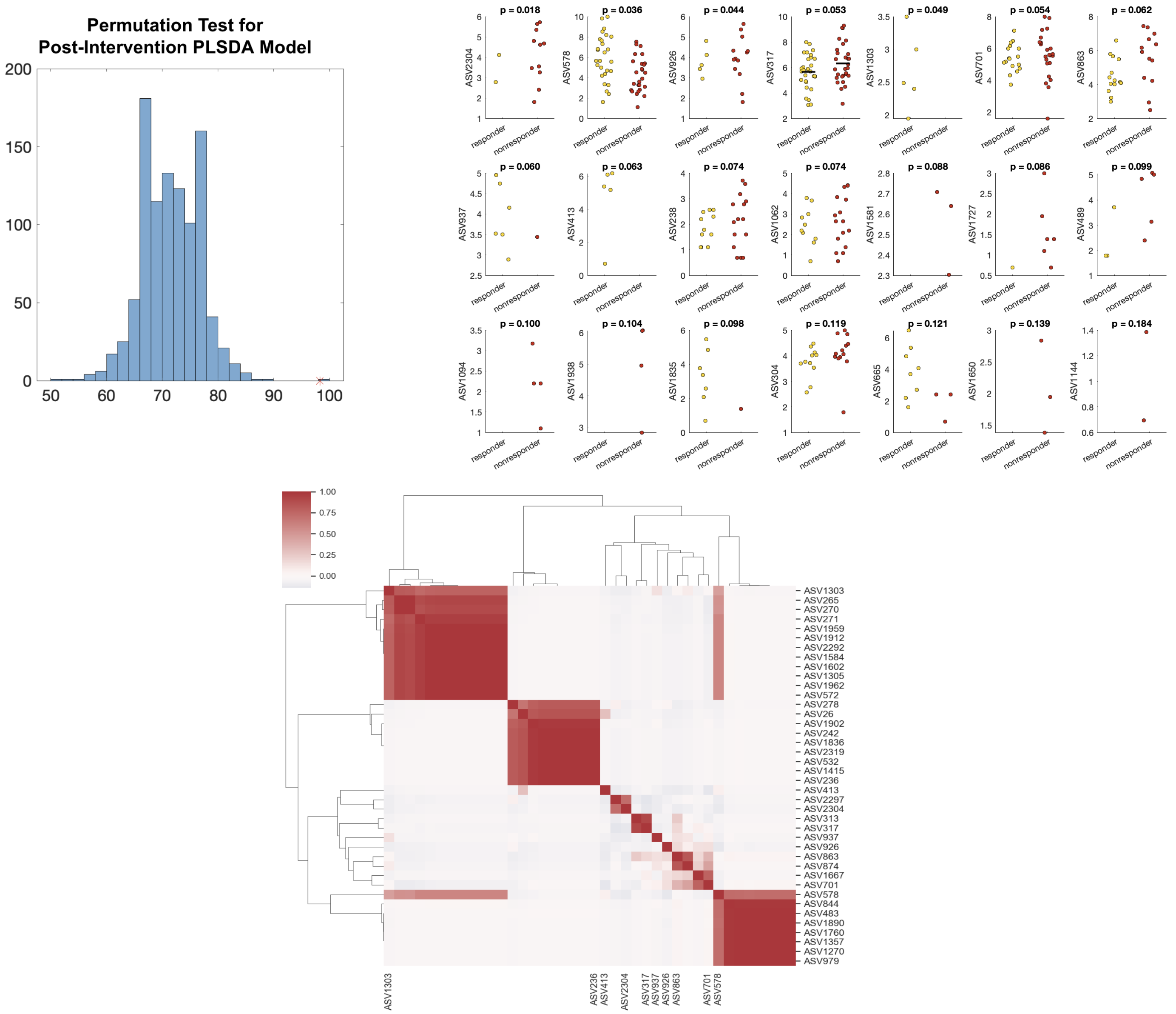


**Fig S6: Additional OPLS-DA figures for Fig. 7 (A)** Results of permutation testing for OPLSDA model in figure 5. The plot shows a histogram of cross-validation accuracy for 1000 randomly permuted models. The correctly labeled model is shown as a red star. **(B)** Jitter plots showing ASV counts per sample for VIPs identified in OPLSDA analysis. **(C)** Correlation network of ASVs with >70% correlation of ASV identified in OPLS-DA. The X-axis has only important features labeled, while the y-axis has correlated and important features labeled.

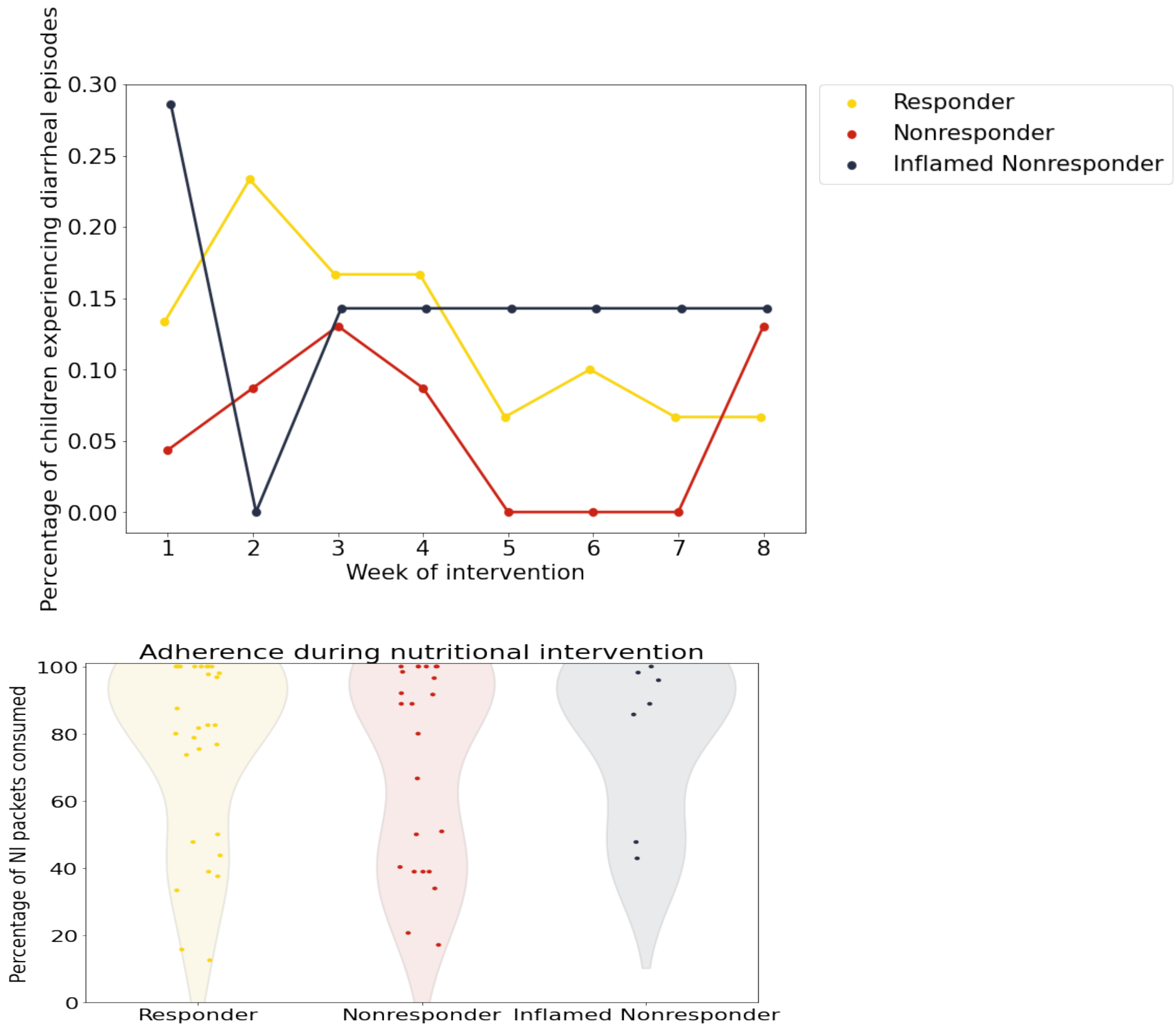


**Fig S7: Inflamed nonresponders show similar patterns of adherence and diarrheal episodes but increased respiratory infections: (A)** Mean number of diarrheal episodes per week of the intervention in each group, error bars show standard deviation. **(B)** Adherence to nutritional intervention by group as shown by the percentage of Acha Mum packets consumed out of total packets distributed for each child during the intervention

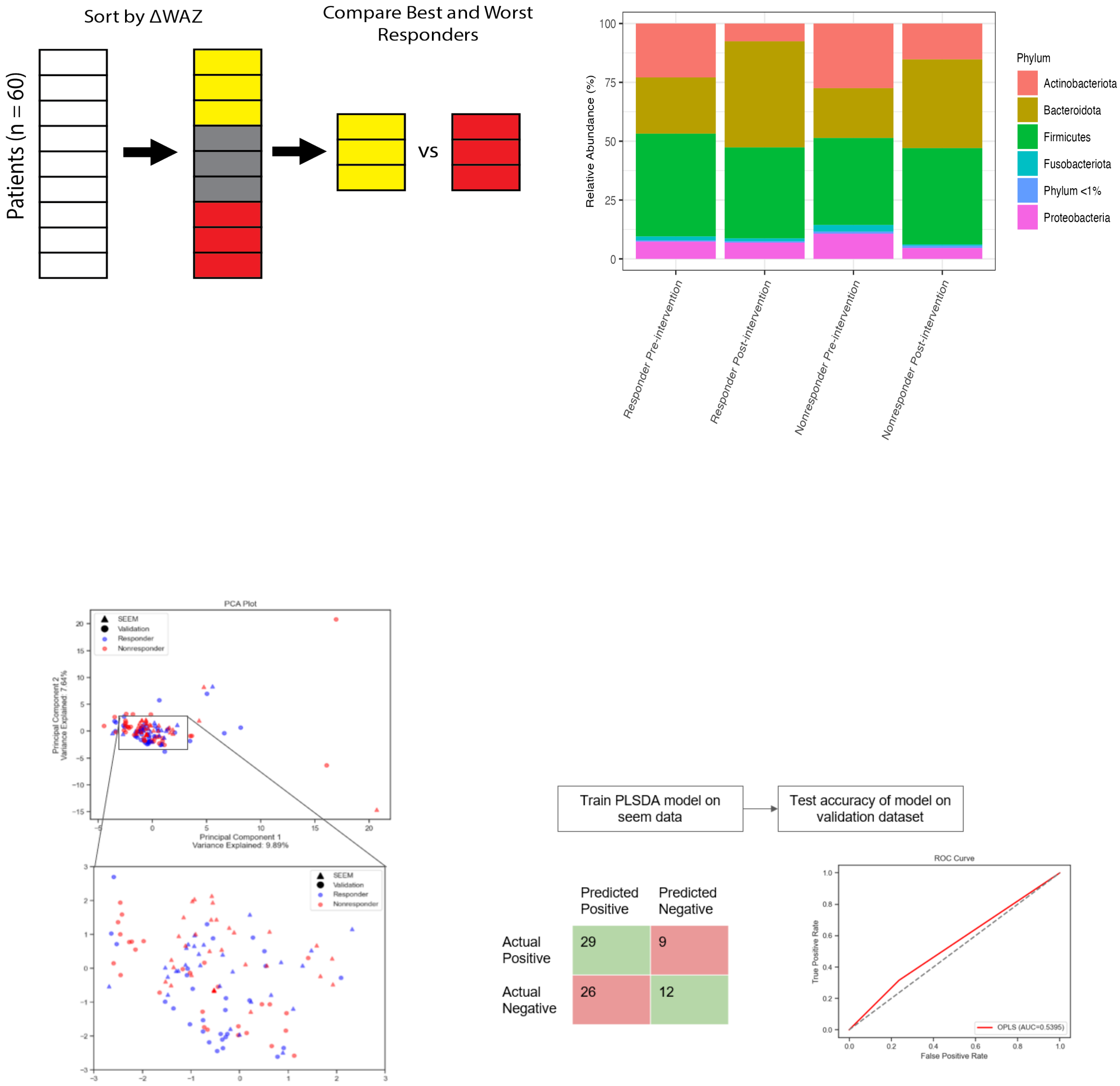


**Fig S8: Separate cohort of malnourished children shows different trends in microbiome restructuring during the intervention: (A)** Children from the validation dataset who received the chickpea-based intervention were sorted by change in WAZ during the intervention, and the 20 best and 20 worst responders were compared. **(B)** Bar plots of microbiome composition at pre and post-intervention timepoints in responder and nonresponder groups. **(C)** Scatter plot of the X scores on latent variables 1 and 2 (LV1 & LV2), where each point represents one sample. **(D)** The PLS model was trained on SEEM data and tested on Chen et al. data. Confusion matrix and ROC curves describe the accuracy of the model.
